# Breast cancer multigene germline panel testing in mainstream oncology based on clinical-public health utility (cancer mortality benefit): ESMO Precision Oncology Working Group recommendations

**DOI:** 10.1101/2025.02.25.25322856

**Authors:** C. Turnbull, M.I. Achatz, J. Balmaña, E. Castro, G. Curigliano, C. Cybulski, S.M. Domchek, D.G. Evans, H. Hanson, N. Hoogerbrugge, P.A. James, A. Krause, K. L. Nathanson, J. Ngeow Yuen Yie, M. Robson, M. Tischkowitz, B. Westphalen, W.D. Foulkes

**Affiliations:** Division of Genetics and Epidemiology, The Institute of Cancer Research, London, UK; Oncology Center, Hospital Sírio-Libanês, São Paulo, Brazil; Hereditary Cancer Genetics Group, Vall d’Hebron Institute of Oncology (VHIO), Barcelona, Spain; Medical Oncology Department, Hospital Universitario Vall d’Hebron, Vall d’Hebron Hospital Campus, Barcelona, Spain; Department of Medical Oncology, Hospital Universitario 12 de Octubre, Instituto de Investigación Sanitaria Hospital 12 de Octubre, Madrid, Spain; Division of New Drugs and Early Drug Development for Innovative Therapies, European Institute of Oncology, IRCCS, Milan, Italy; Department of Oncology and Hemato-Oncology, University of Milan, Milan, Italy; International Hereditary Cancer Center, Department of Genetics and Pathology, Pomeranian Medical University in Szczecin, Szczecin, Poland; Divisions of Hematology/Oncology, Department of Medicine, Perelman School of Medicine, University of Pennsylvania, Philadelphia, PA, USA; Basser Center for BRCA, Abramson Cancer Center, University of Pennsylvania, Philadelphia, PA, USA; Manchester Academic Health Science Centre, Division of Evolution and Genomic Sciences, School of Biological Sciences, Faculty of Biology, Medicine and Health, The University of Manchester, Manchester, UK; Department of Clinical and Biomedical Sciences, University of Exeter Medical School, Exeter, United Kingdom; Peninsula Regional Genetics Service, Royal Devon University Healthcare NHS Foundation Trust, Exeter, United Kingdom; Department of Human Genetics, Radboud University Medical Center, Nijmegen, The Netherlands; Parkville Familial Cancer Centre, The Royal Melbourne Hospital and Peter MacCallum Cancer Centre, Melbourne, Victoria, Australia; Division of Human Genetics, National Health Laboratory Service and School of Pathology, The University of the Witwatersrand, Johannesburg, South Africa; Division of Translational Medicine and Human Genetics, Department of Medicine, Perelman School of Medicine, University of Pennsylvania, Philadelphia, PA, USA; Cancer Genetics Service, Division of Medical Oncology, National Cancer Centre Singapore, Singapore; Lee Kong Chian School of Medicine, Nanyang Technological University, Singapore; Department of Medicine, Memorial Sloan Kettering Cancer Center, New York, New York, USA; Department of Medicine, Weill Cornell Medical College, New York, New York, USA; Department of Medical Genetics, National Institute for Health Research Cambridge Biomedical Research Centre, University of Cambridge, Cambridge, United Kingdom; Comprehensive Cancer Center Munich & Department of Medicine III, Ludwig-Maximilian University of Munich, Munich, Germany; German Cancer Consortium (DKTK Partner Site Munich), Heidelberg, Germany; Departments of Human Genetics, Oncology and Medicine, McGill University, Montréal, QC, Canada

## Abstract

**Background:** With widening therapeutic indications, germline genetic testing is offered to an increasing proportion of patients with breast cancer (BC) via mainstream oncology services. However, the gene set tested varies widely from just BRCA1/BRCA2 through to ‘pan-cancer’ panels of near 100 genes. If a germline pathogenic variant (GPV) is detected, the BC proband and other family GPV-carriers may be offered interventions such as risk-reducing surgery and decades of intensive surveillance for the various cancers linked to that gene.

**Methods:** ESMO’s Precision Oncology Working Group established an international expert group in breast cancer germline genetics. This group firstly established a framework of criteria by which to evaluate each breast cancer susceptibility gene (BCSG) for inclusion on a breast cancer multigene panel test (BC-MGPT) for universal mainstream testing for BC cases. Next the panel scored BCSGs for gene utility regarding (i) BC risk estimation, (ii) clinical actionability (iii) evidence of impact on cancer-specific mortality (and/or morbidity).

**Results:** The group agreed genes should be included on the BC-MGPT based on potential cancer-specific mortality (and/or morbidity) benefit. Judged as of high or moderate utility on this basis were 7 genes: *BRCA1, BRCA2, PALB2, RAD51C, RAD51D* and *TP53* (for BC diagnosed <40 years), with *BRIP1* later added. Whilst potentially informative for BC risk estimation, *CHEK2* and *ATM* were judged to offer insufficient evidence for improving cancer-specific mortality. The expert group recommended strongly against inclusion of ‘syndromic’ genes such as *STK11, PTEN, NF1* and *CDH1*.

**Conclusion:** With expanded germline testing in patients with BC (and cascade testing into families), the number and nature of resultant GPV carriers identified will be dictated by the genes included on the upfront BC-MGPT. The potential harms, opportunity and economic costs of decades of surveillance of multiple organs and risk-reducing surgeries should be outweighed by strong evidence of meaningful benefit, improved cancer-specific mortality (and/or morbidity).

**Highlights:**

- ESMO expert panel settled a list of genes for inclusion on a BC-MGPT based on potential cancer-specific mortality benefit
- This BC-MGPT should include 7 genes: *BRCA1, BRCA2, PALB2, RAD51C, RAD51D, BRIP1* and *TP53* (for BC diagnosed <40 years)
- This BC-MGPT would service urgent diagnostic mainstreaming germline testing requirements for all eligible BC cases
- ‘Syndromic’ genes such as *STK11, PTEN, NF1* and *CDH1* should only be tested downstream post expert review in a minority of BC
- The mortality benefit was deemed equivocal for *ATM* and *CHEK2*, being primarily of intermediate penetrance for ER-positive BC

## Introduction

The only breast cancer susceptibility genes (BCSGs) that had been identified by the turn of the century were those of sufficiently high penetrance to have generated a linkage signal^1,2^. Clinical genetic testing was expensive, low throughput and accordingly restricted to a small number of families with multiple cases of breast and ovarian cancers (for *BRCA1* and *BRCA2*) or characteristic syndromic features (for genes such as *PTEN* and *STK11*). Clinical testing was likewise restricted to just the specific gene(s) most likely to ‘confirm’ the suspected diagnosis already manifest in the patient or family. Predictive testing of the germline pathogenic variant (GPV) was then offered to family members to provide a dichotomous result of their being either at very high risk of or spared from the familial predisposition.

Subsequent linkage analyses in hereditary breast-ovarian cancer (HBOC) families were unfruitful, confirming there to be no additional genes with a risk-frequency profile comparable to *BRCA1*/*BRCA2*^*3*^. With accordant shift to case-control studies of genes in candidate pathways (often related to DNA repair), association with breast cancer has over the following two decades been reported for multiple additional genes. *PALB2, CHEK2* and *ATM* were established early as BCSGs with risks being (reasonably) reproducible over time^4–8^. By contrast, only with large population-based case-control studies (and meta-analysis thereof) have we been able to confirm association of GPVs in *BARD1, RAD51C*, and *RAD51D* with breast cancer (BC), with the rarity of GPVs and the subtype-specific association with triple negative breast cancer (TNBC) contributing to inconsistency of earlier results^9–11^. These large population-based case-control studies have also been sufficiently well-powered to refute a multitude of other previously reported BC associations, including genes involved in DNA repair pathways (for example *BRIP1, NBN, RAD50, RECQL, XRCC2, XRCC3, SLX4* and *GEN1*) and mismatch repair genes^12–16^. Notably, for *TP53, PTEN, STK11* and *CDH1*, BCSGs identified via their distinctive syndromes of pleomorphic susceptibility to rare cancers (hereafter termed “syndromic BCSGs”), the association signals in these population-based breast cancer case-control studies were markedly lower than those from earlier familial-based estimates.

The initial identification of *BRCA1* and *BRCA2* was based on epidemiologically observed co-susceptibility for breast cancer *and* ovarian cancer (OC). Additional studies in BRCA1/BRCA2 families have suggested association with prostate and pancreatic cancers; relative risks are of more modest magnitude than for breast and ovarian cancer but seem largely reproducible for BRCA2. The other BCSGs with BRCA1/BRCA2-related DNA repair candidacy (*PALB2, ATM, CHEK2, RAD51C, RAD51D, BARD1*) have thus been extensively studied for association with these cancers, with varying reproducibility of findings) and have been grouped under the umbrella of HBOPP (hereditary breast-ovarian-prostate-pancreatic cancer) genes. Numerous associations with other rare and common cancers have been reported on studies of both the HBOPP and syndromic BCSGs.

Clinical management protocols for GPV carriers of these genes have been evolved, aimed at mitigating the elevated risks for of breast and other implicated cancers via SPED (surveillance, prevention, early detection) interventions. These surveillance protocols for GPV carriers typically (i) are greater frequency, (ii) are initiated at younger age and continue for a longer duration (many decades) and (iii) involve multi-modal approaches (e.g. combining serum biomarker testing with multiple imaging modalities), as compared to the population-level screening recommended for the given cancer type (if any). Whilst programmes of population-level screening are typically only implemented following randomised trials demonstrating cancer-specific mortality benefit (quantified against false-positive and overdiagnosis rates), the intensive surveillance recommendations for PV-carriers are often reliant on expert opinion or modelling due to lack of direct evidence from longitudinal clinical studies (let alone randomised trials).

Over the last decade, embedding of Next Generation Sequencing (NGS) within diagnostic laboratories now means that the wet-laboratory costs for testing of 200 genes differ little from those for 20 or 2 genes. This technological economy has catalysed a dramatic shift in clinical testing practice. Previously, expensive, low throughput testing necessitated that clinicians undertake individualised selection of the gene(s) for testing based on patient phenotype and clinical context (even, for example, strategic sequencing of *BRCA2* ahead of *BRCA1*). Now we typically “tick-box order” a routine pre-designed ‘panel’ of genes relevant to breast cancer susceptibility, a so-called breast cancer multi-gene panel test (BC-MGPT). The number and range of genes included on this BC-MGPT varies widely between countries and providers (varying from 7-37 genes) (Table 1). Depending on local protocols and reimbursement, a newly diagnosed BC patient may instead be offered a pan-cancer panel (typically >50 genes) to obviate downstream testing requirements.

**Table 1:**
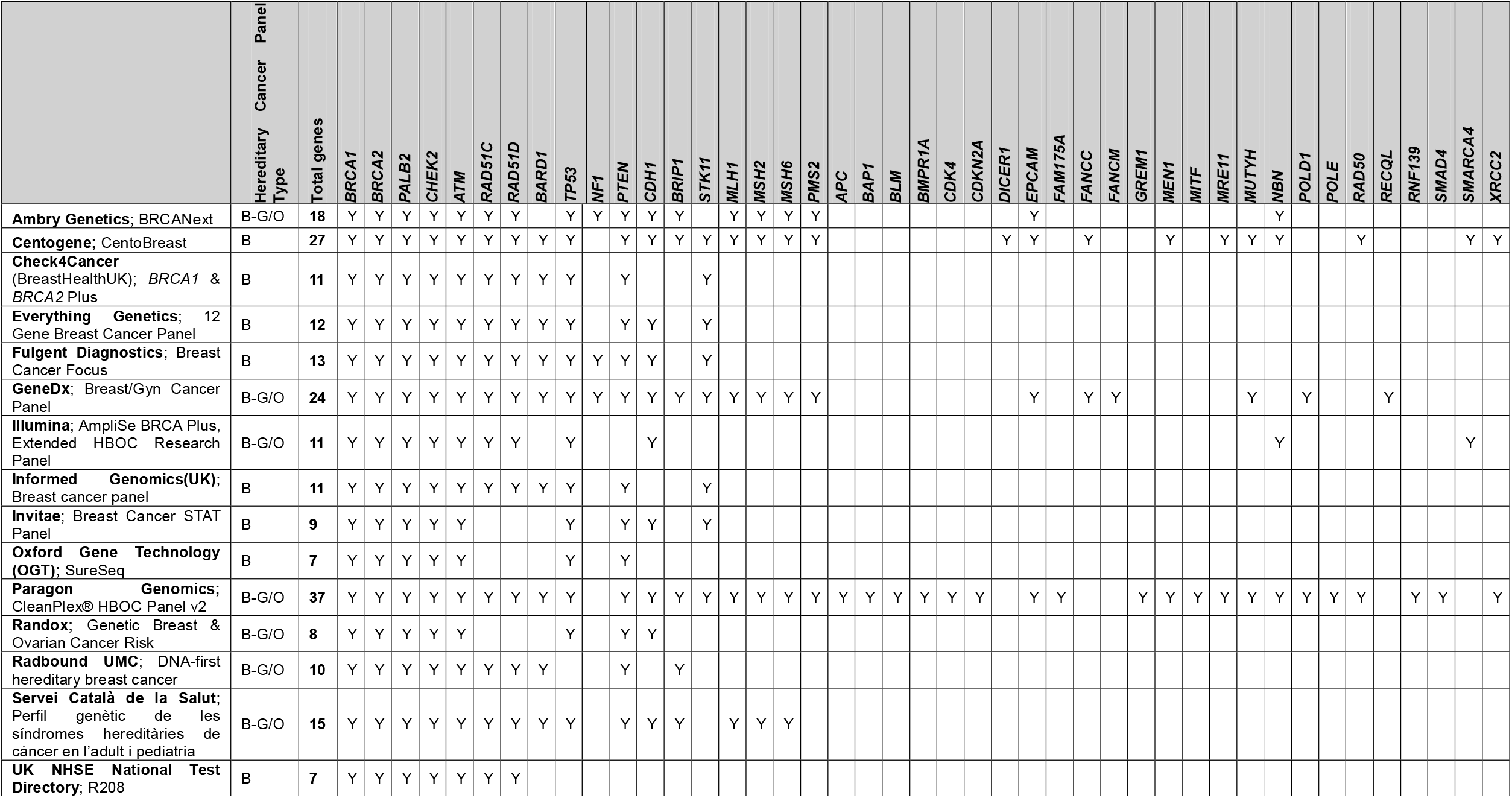
Survey of genes included on 12 commercial and 3 health-service multi-gene panel tests designated for breast cancer (+/−ovarian/gynae cancers) B: breast, B-G/O: breast plus gynaecological/ovarian

Extended therapeutic indications for PARP inhibitors (PARPi) have increased the proportion of patients with BC offered germline testing, adding momentum to arguments for broader or even universal germline testing for all female BC presentations. Indeed, the American Society of Clinical Oncology (ASCO) recently recommended that germline genetic testing is offered routinely to all female BC diagnoses age <65 (and many others)^17^.

However, as an increasing proportion of BC cases are tested upfront, more GPVs will be detected in the probands. In turn, there will be more relatives with GPVs identified on familial cascading. In turn, there will be commensurate expansion in the volume of SPED interventions offered to probands and their relatives. The resultant volume of GPV carriers identified, the scale of concomitant SPED activity and the overall impact on cancer mortality will be directly predicated on the number and specifics of the genes included upfront in the BC proband panel.

**Box 1: Measures of Clinical Utility** for evaluation of genes to be included on a breast cancer multigene germline panel test (BC-MGPT)

**1) Clinical Validity:** Is there a robust reproduceable association of gene GPVs with breast cancer? If so. the GPV is deemed of utility this finding contributes towards BC risk estimation

**2) Clinical Actionability:** Are there society-or national clinical guidelines advising SPED interventions for cancers reported to be associated with GPVs in this gene? If so. the GPV is deemed of utility in having “clinical actionability” (that the GPV “alters clinical management”).

**3) Cancer-specific mortality benefit**\*: Is there evidence that the oncological interventions and/or SPED interventions implemented in GPV-carriers reduce the likelihood of dying from the corresponding cancers? If so, the GPV is deemed to confer “net clinical-public health” utility.

* All-cause mortality impact (thus including potential off-target harms and competing risks) would typically be deemed a more rigorous metric of benefit

**Net clinical-public health utility would also encompass, where available, evidence of Quality of Life (morbidity) benefit

## Aims

The expert group was convened at the recommendation of the ESMO Precision Oncology Working Group, via individual invitations based on expertise and representation across healthcare settings. The overarching aim for the expert group was to consider which genes should be included on a breast cancer multigene germline panel test (BC-MGPT), specifically relating to incipient expansion of germline genetic testing in patients with BC relating to expanded therapeutic indications.

The expert group established in early discussions that the criteria by which genes are currently included for germline testing, as per Box 1, are (i) **clinical validity** (i.e. GPV finding informs BC risk estimation on basis of robust evidence of association) and (ii) **clinical actionability** (i.e existence of SPED recommendations for associated cancers). The expert group agreed that whilst these two parameters had broadly been taken to be proxies for improved survival (mortality), available evidence was often lacking or contradictory to this assumption. The expert group thus agreed as a first aim to consider genes for inclusion on a BC-MGPT for their utility regarding each of these three measures (i) breast cancer risk estimation (ii) actionability for SPED interventions for associated cancers (iii) improving cancer-related mortality.

The expert group thus sought to define more clearly the “routine-mainstream” BC probands for whom our recommended BC-MGPT would be applicable. The expert group recognised that eligibility criteria for BC-MGPT relating to age, histology/characteristics of the breast cancer and/or personal and/or cancer family history are implemented in most healthcare settings, but that these vary widely. The expert group agreed that we should seek to define a single, universal BC-MGPT that would be applicable for (virtually) all eligible BC cases, meaning that:

i. the BC-MGPT would be applicable regardless of the healthcare settings and of threshold of eligibility. Hence, the panel would be appropriate for all eligible patients with BC (a) in healthcare settings in which eligibility is currently more restricted (eg~20% of BC cases in UK, or other settings where lower) (b) in healthcare settings in which eligibility is currently more permissive (e.g., ~40-50% of BC cases eligible via ASCO criteria) or even (c) if eligibility were extended to all BC cases.
ii. the genes on the BC-MGPT would be a sufficient test for >99% of the eligible patients with BC. The test would thus be readily implementable in the urgent diagnostic mainstream oncology (without requiring additional algorithms of individualised patient assessment to determine gene selection).
iii. additional genetic assessment would only be indicated in a tiny minority (<1%) of BC cases. These could be identified by questionnaire downstream of routine early BC-MGPT and referred to clinical genetics.

We thus use the term “routine-mainstream” BC probands to describe recently diagnosed patients with BC being offered this BC-MGPT in a mainstream oncology setting (encompassing settings of stricter, looser or no eligibility restrictions).

The expert group agreed that in the context of (i) incipient expansion of germline genetic testing in BC and (ii) the scale of SPED activity across a family consequent from a GPV being identified in the proband, greater focus was required regarding evidenced impact on cancer-related mortality. The expert group thus agreed a second aim to make explicit recommendations regarding genes for inclusion on a BC-MGPT, based on evaluation of impact on cancer mortality (hereafter termed clinical-public health utility). Namely that the BC-MGPT should include genes on the basis of available evidence that (i) associated cancer risks and concomitant SPED interventions number will result in improvement to cancer mortality for GPV carriers and/or (ii) systemic oncology management offers survival benefit in the proband.

### Objectives

The expert group established four objectives by which to address their aims:

1. To review genes currently included on BC-MGPTs-to identify a set of genes for review
2. To define and assemble evidence relevant for per-gene evaluations
3. To undertake individual evaluations of each gene against a range of different measures
4. To integrate individual-level evaluations to provide consensus recommendations regarding genes for inclusion for a BC-MGPT applicable for “routine-mainstream” BC cases in mainstream oncology, as judged by clinical-public-health-utility (evidenced impact on cancer mortality)

## Methods

We convened four meetings of the expert group, with development of recommendations based on quantitative polls conducted between the meetings (see Supplementary Methods).

### Outcomes from Objectives

#### Objective 1: To review genes currently included on BC-MGPTs

The expert group identified eleven BC-MGPTs offered by commercial providers and three from health-services which were specified for either (i) BC (ii) breast and/or ovarian cancer or (iii) breast and/or gynae cancers (Table 1). Through this review, the expert group agreed a core set of 13 widely included BCSGs for evaluation, which we grouped into two broad categories:

**1) HBOPP-related genes** *BRCA1, BRCA2, PALB2, ATM, CHEK2, RAD51C, RAD51D* and *BARD1*. These genes are grouped together on the basis of (i) being identified through common pathways related to DNA-repair and/or (ii) broad commonality of spectrum of additional reported cancer associations (potentially including ovarian, prostate and/or pancreatic cancer). *BRIP1*, an ovarian-only susceptibility gene, was later included by the expert group for consideration.

**2) Rare ‘syndromic’ genes** *TP53, CDH1, NF1, STK11* and *PTEN*. These genes are grouped together on the basis of each causing a rare cancer-susceptibility syndrome for which non-malignant features would typically precede cancer onset (*NF1, STK11* and *PTEN*) and/or the constellation of rare cancers is typically distinctive (*TP53, CDH1*).

#### Objective 2: To assemble evidence for consideration in per-gene evaluations

The expert group agreed five categories of evidence for consideration within the per-gene evaluations of utility (for additional details of expert group review see Table 2 and Appendix Tables 1-4).

**Table 2:** Frequency of pathogenic variants, cancer associations and efficacy of gene-stratified systemic oncological for 13 Breast Cancer Susceptibility Genes: Associated cancers reported as per expert group consensus (bold: well established; non-bold: less well established; brackets: reported but equivocated; see Appendix Table 5 for source references. Other Fields (⍰⍰⍰⍰⍰: highest); scored as per expert group consensus

|  | Evaluation of in unselected breast cancer<br>(meta-analysis of BRIDGES, CARRIERS, UKBIOBANK) |  |  |  |  | Ovarian<br>cancer<br>association | Mortality<br>benefit:<br>systemic<br>oncological<br>manageme<br>nt (stratified<br>by gene<br>GPV) | Other cancer associations (non breast/ovarian) |  |  |
| --- | --- | --- | --- | --- | --- | --- | --- | --- | --- | --- |
|  | Frequency of Pathogenic<br>variants |  | Association of GPVs with breast<br>cancer |  |  |  |  | Tumours reported as associated | Instability<br>between<br>studies for<br>reported<br>associations | Uncertainty of<br>risk estimates<br>due to<br>ascertainment |
|  | In unselected<br>breast cancer<br>cases | In the control<br>population | Overall odds<br>ratio for breast<br>cancer | Triple<br>negative BC<br>enrichment | ER-positive<br>BC<br>enrichment |  |  |  |  |  |
| **BRCA1** | 0.99%<br>1 in 101 | 0.088%<br>1 in 1,134 | 8.73<br>(7.47-10.20) | ████ |  | ██████ | ██████ | *Prostate, stomach, pancreas, male breast* | ██ | █ |
| **BRCA2** | 1.48%<br>1 in 68 | 0.25%<br>1 in 400 | 5.68<br>(5.13-6.30) | █ |  | ████ | ████ | **Prostate, pancreas, male breast, stomach, (melanoma)** | ███ | █ |
| **PALB2** | 0.54%<br>1 in 187 | 0.14%<br>1 in 726 | 4.30<br>(3.68-5.03) | ██ |  | ██ | █ | **Pancreas, male breast** | ██ | █ |
| **CHEK2** | 1.36%<br>1 in 73 | 0.58%<br>1 in 172 | 2.40<br>(2.21-2.62) |  | ████ |  |  | **Prostate, kidney, thyroid, (osteosarcoma, colorectal)** | ████ | █ |
| **ATM** | 0.76%<br>1 in 132 | 0.35%<br>1 in 287 | 2.16<br>(1.93-2.41) |  | ████ | █ |  | **Prostate, pancreas** | ████ | █ |
| **BARD1** | 0.15%<br>1 in 672 | 0.062%<br>1 in 1,621 | 2.34<br>(1.85-2.97) | ███ |  |  |  | [nil of note] |  | █ |
| **RAD51C** | 0.11%<br>1 in 913 | 0.055%<br>1 in 1,819 | 1.53<br>(1.15-2.04) | ███ |  | ███ |  | [nil of note] |  | █ |
| **RAD51D** | 0.093%<br>1 in 1,079 | 0.048%<br>1 in 2,100 | 1.76<br>(1.15-2.41) | ███ |  | ███ |  | [nil of note] |  | █ |
| **TP53** | 0.054%<br>1 in 1,844 | 0.0073%<br>1 in 13,606 | 3.62<br>(1.98-6.61) |  |  | █ |  | **Sarcomas, adrenocortical carcinoma, neurological , epithelial** | ███ | █████ |
| **PTEN** | 0.027%<br>1 in 3,755 | 0.0067%<br>1 in 14,902 | 2.63<br>(1.38-5.02) |  |  |  |  | **Thyroid, kidney, endometrial, colorectal, hamartomas** | ███ | ██████ |
| **NF1** | 0.068%<br>1 in 1,470 | 0.033%<br>1 in 3,068 | 2.01<br>(1.43-2.83) |  |  |  |  | **Neurofibromas, neurological (mainly benign)** | ██ | ██████ |
| **STK11** | 0.0087%<br>1 in 11,525 | 0.0019%<br>1 in 52,157 | 1.60*<br>(0.48-5.30) |  |  |  |  | **Hamartomas, gonadal, pancreas, other epithelial** | ███ | ██████ |
| **CDH1** | 0.037%<br>1 in 2,668 | 0.015%<br>1 in 6,658 | 2.01<br>(1.25-3.24) |  |  |  |  | **Diffuse gastric cancer** | █ | ██████ |

##### i. Frequency of GPVs in unselected (population-type) breast cancer and magnitude of BC association

The expert group surveyed GPV-frequencies for the thirteen genes in unselected (population-type) BC versus population controls (UK Biobank, BRIDGES, CARRIERS, Table 2). The expert group noted for *BRCA1* and *BRCA2* high odds ratios (ORs) for BC association with enrichment for TNBC and younger-onset disease (in particular for *BRCA1*). The expert group recognised a high but lesser BC risk for *PALB2* (OR 4-5) with some enrichment for TNBC. The expert group noted *ATM* and *CHEK2* to have moderate association with BC (OR 2-3), with risk largely restricted to ER-positive disease. The expert group noted low GPV frequencies and very modest metrics for association in unselected BC cases for *RAD51C* and *RAD51C* (OR<2), albeit with enrichment for TNBC.

The expert group noted that for the ‘syndromic’ BCSGs, the frequency of GPVs in unselected BC cases were very low and only modestly greater than that in the population, resulting in quite modest ORs for association. However, whilst these population-based BC studies are spared the upward-bias in association estimates inherent to familial, phenotype-based ascertainment, these BC series may be depleted for precisely the atypical and/or very young-onset types of BC cases characteristically linked to these genes^18^.

The expert group agreed that whilst a very low GPV-frequency in unselected (population-type) BC should not preclude inclusion of the gene on the BC-MGPT. The expert group nevertheless noted that where the rate true GPVs was very low, there would a proportionately higher ratio of VUSs, with implications for complex, time-consuming variant interpretation, challenging clinical management and patient anxiety.

##### ii. Per-gene associations with other cancers

Co-aggregation of breast and ovarian cancers had been empirically observed long before linkage analyses in such families, which enabled identification of *BRCA1* and *BRCA2* and quantitation of commensurately high penetrance (and reproducible) estimates for these cancer types. For *BRCA1* the risk to age 80 for developing OC is estimated at ~44% (SIR~50) and for *BRCA2* ~17% (SIR~14)^19^,

Studies associating *BRCA1*/*BRCA2* with other cancer types, as well as those investigating cancer associations for other BCSGs, have been more challenging. Studies based on familial/phenotypic ascertainment may suffer upwardly biased estimates of effect-size; there may also be inflated cancer incidences against background rates where extra surveillance has been implemented. Early case-control studies were often distorted due to differential molecular analyses for controls versus cases with added variability due to case definition and the spectrum of variants included. However, overarching these methodological limitations is the impact of statistical variation where (i) effect-sizes are modest, (ii) GPVs are comparatively rare and/or (iii) the cancer type is of low population frequency and/or late onset.

The expert group noted robust reproducible associations with ovarian cancer (OC) for *RAD51C* and *RAD51D* (OR 6-9; risk to age 80 to 11-13%)^20,21^. However, for other HBOC genes, association with OC is evidently of more modest magnitude with consequently less reproducibility for reported gene associations across studies. *PALB2* and *ATM* have been more consistently associated with OC with risks seemingly 2-4-fold elevated (Table 2, Appendix Tables 3, 4)^20,22-27^.

**Table 3:**
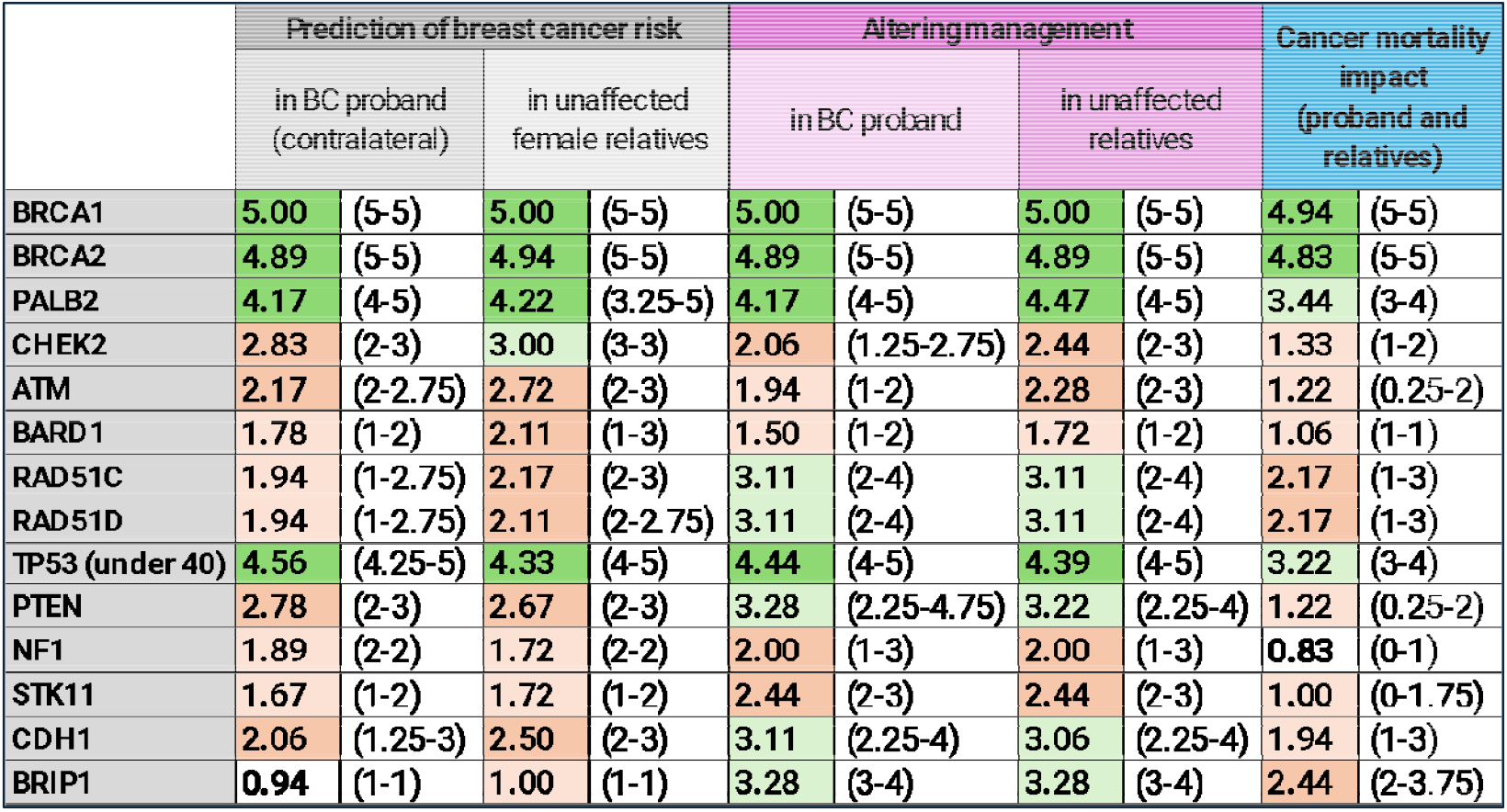
Mean and interquartile range of individual scores from 18 expert group members for five measures of gene utility for 14 genes. The utility relates to identification of a GPV on testing of the gene in “mainstream-routine” breast cancer proband. Mean scores are presented to 1 dp: 4-5 (very high, *dark green*), 3-3.99 (high, *pale green*), 2-2.99 (moderate, *dark orange*), 1-1.99 (low, *pale orange*), 0-0.99 (very low, uncoloured). *TP53* was scored specifically in the context of testing only BC probands diagnosed <40 years.

In studies of *BRCA1/BRCA2* families, elevated risk of male breast, prostate and pancreatic cancers in *BRCA2* families have been the most reproducible of the multitude of cancer associations reported. Recent prospective analyses in *BRCA2* GPV carriers from the international CIMBA consortium estimate risk to age 80 at 2.5% for pancreatic cancer (RR = 3.34; 95% CI, 2.21 to 5.06) and at 27% for prostate cancer (RR = 2.22; 95% CI, 1.63 to 3.03)^28^. Because of their links to *BRCA1*/*BRCA2*, pancreatic and prostate cancers have been a particular focus of study for the other HBOC genes. Although with considerable variability between studies, the expert group noted relatively consistent reported association with pancreatic cancer for *ATM* GPVs (OR/RR 4-8), with less reproducible estimates of association for *PALB2*^25,29-32^. Albeit with widely varying estimates, prostate cancer has also been relatively consistently reported for *ATM* and *CHEK2*, with recent analysis in UK Biobank suggesting respective ORs of 2.35 (1.78 to 3.11) and 1.92 (1.59 to 2.32)^25^.

For rare cancer types linked to highly rare ‘syndromic’ susceptibility genes, case-control studies of sufficient magnitude using unselected case series are not typically feasible. Accordingly, for the ‘syndromic’ BCSGs working risk estimates for the rare cancers implicated in the syndromic spectrum are typically still based on phenotype-driven ascertainment of families. Whilst studies suggest such risks may be substantially upwardly biased^33^, the expert group agreed there to be great uncertainty regarding true prospective risks of rarer cancers when a is GPV identified outside of the characteristic personal/familial phenotype.

##### iii. *Evidence of cancer-specific* mortality benefit *from available SPED interventions*

The expert group noted that where there was direct evidence from longitudinal (albeit largely non-randomised) clinical studies, these data typically related to carriers of BRCA1/BRCA2 GPVs only. Thus, the expert group noted requirement for extrapolation of these data to PV carriers of other genes, necessarily taking into account differences in cancer risk and subtype associations.

The expert group noted several studies demonstrating ovarian cancer-specific mortality benefit for RRBSO in *BRCA1/BRCA2* GPV-carriers, with benefit in breast-cancer-specific mortality and all-cause mortality also reported in some studies ^34–38^. The expert group noted absence of studies investigating mortality benefit from RRBSO for genes such as *RAD51C/RAD51D/BRIP1*. Based on (i) the sizeable OC-specific mortality benefit from RRBSO for *BRCA1/BRCA2* GPV-carriers (ii) modelling analyses that this mortality benefit from RRBSO would extend to those with a lifetime risk of OC of ≥5% (iii) estimated lifetime OC risk of ~10% for females with RAD51C/RAD51D/BRIP1 GPVs, the expert group concurred that mortality benefit from RRBSO could reasonably be extrapolated to *RAD51C, RAD51D* and *BRIP1* GPV-carriers (albeit with a commensurately lower predicted impact on mortality) ^39,40^.

The expert group noted recently-published internationally-amalgamated follow-up studies reporting survival benefit from breast MRI imaging in unaffected *BRCA1* GPV carriers, noting the absolute benefit in this study to be modest and results from previous studies to be equivocal^41^. The expert group also noted a comparison study reporting a small absolute survival benefit for RRBM over MRI surveillance in unaffected *BRCA1* carriers. Studies of RRBM have consistently reported a dramatic reduction in the incidence of BC but have typically reported low numbers of events (deaths) meaning in many studies the incidence reduction has not translated into mortality benefit; treatment-related improvements in BC outcomes over the last two decades necessarily will have extended the follow-up required to evaluate benefit from RRBM^42–45^. The expert group overall agreed that data supported RRBM and MRI as conferring some modest BC-specific mortality for *BRCA1* GPV carriers. The expert group noted disparity between *BRCA1* and *BRCA2*-PV carriers in mortality benefit, the expert group were of mixed opinion regarding the confidence with which mortality benefit could be extrapolated from *BRCA1* to other *GPV* carriers such as *TP53* and *PALB2*.

The expert group noted the lack of direct evidence quantifying mortality impact in unaffected *CHEK2*/*ATM* GPV carriers for enhanced MRI surveillance or RRBM. Based on (i) data for *BRCA2* versus *BRCA1*, (ii) lower BC-risks of *CHEK2/ATM* compared to *BRCA2*, (iii) predominant association of *CHEK2/ATM* with ER-positive BC, (iv) recent modelling analyses, the expert group concurred any absolute breast cancer-specific mortality benefit (above routine breast screening) for enhanced breast surveillance or RRBM in those with *CHEK2/ATM* GPVs was likely at best to be extremely modest^46^.

The expert group noted that for the affected BC proband, survival was primarily predicated on the outcome from her first BC rather than SPED interventions for the contralateral breast. This has demonstrated for *BRCA1*/*BRCA2* GPV carriers; the expert group concurred could reasonably be extrapolated to *CHEK2/ATM* GPV-carriers (where BC risks are lower and association is with ER-positive BC). The expert group noted the wide variation in reported estimates of elevation in contralateral breast cancer risk (if any) associated with PVs in these *CHEK2/ATM*^47–49^.

The expert group noted that public-health bodies such as the United States Preventive Services Task Force (USPSTF) and UK National Screening Committee (UK-NSC) currently recommend against screening at population-level for either prostate cancer (due to excessive over-diagnosis) or for pancreatic cancer (due to lack of proven impact on mortality of available technologies)^50,51^. The expert group noted that whilst regimens of surveillance for pancreatic and prostate cancer are widely implemented for GPV-carriers of various BCSGs, evidence is currently lacking as to whether these surveillance regimens confer any net benefit in cancer mortality and/or adequate mitigation of cancer overdiagnosis in these groups^52–54^.

The expert group noted various intensive protocols for multi-organ system surveillance implemented for individuals with GPVs in ‘syndromic’ BCSGs. The expert group noted that such regimens had (unsurprisingly) never been evaluated via randomised trials, meaning it will remain unclear whether perceived survival benefits from these surveillance regimens are due to lead time or genuine improvement in mortality; likewise, it remains impossible to quantify the extent and impact of overdiagnoses from these regimens.

##### iv. Potential physical harms, psychological implications and problems with uncertainty

The expert group noted that mainstream oncology clinicians (and consequently their patients) may lack appreciation of the potential harms of genetic testing and the downstream consequences thereof, in particular (i) the potential physical harms from long-term intensive regimes of surveillance, including overdiagnosis and overtreatment (ii) negative physical, aesthetic and/or psychosexual sequelae from risk-reducing surgery and (iii) the psychological burden of lifelong genetic assignation of elevated cancer risk and opportunity costs of management thereof ^55^. The expert group were in strong agreement that the benefit-harm trade-off varied according to (a) the robustness of cancer risk estimates (b) the clinical context in which the genetic test was performed (c) quality of evidence underpinning the mortality benefits of concomitant SPED interventions^56,57^. The expert group agreed that harms would most commonly outweigh benefits where aggressive surveillance/surgery was implemented for GPV-carriers of ‘syndromic’ BCSGs ascertained in contexts in which cancer risks were highly uncertain. For example, for a GPV in *CDH1* or *STK11* detected in a BC proband in the absence of any manifestation of relevant familial disease and/or syndromic features ^58–61^.

##### v. Evidence for gene-based stratification in systemic oncological management

The expert group noted that clinical trials in the metastatic and adjuvant setting have demonstrated impact on progression-free survival and, in the adjuvant setting, overall survival from use of PARP inhibitors and/or platinum following stratification by *BRCA1/BRCA2* mutational status^62,63^. The expert group noted also demonstration of improved clinical outcomes in the metastatic setting from treatment stratification by *PALB2* mutational status ^64,65^.

#### Objective 3: To generate individual and summary scores for genes for different measures of utility

Following expert group review of the evidence items assembled for Objective 2, the eighteen members of the expert group independently awarded integer scores of 0-5 for each gene for the utility of identifying GPV in the context of testing of “routine-mainstream” BC probands. Thirteen (subsequently fourteen) genes were scored regarding their utility for:

i. estimation of BC risk in the proband (contralateral breast management)
ii. estimation of BC risk in unaffected female relatives
iii. alteration of clinical management (actionability) in the proband
iv. alteration of clinical management (actionability in (unaffected) relatives
v. evidenced mortality benefit in the proband and/or relatives (clinical-public health utility)

The mean and interquartile range for the scores of the eighteen contributing expert group members are presented in Table 3.

#### Objective 4: To generate consensus recommendations for genes to be included on a BC-MGPT

Reflective of the consensus scoring (mean, IQR), the expert group agreed that a BC-MGPT based on evidenced cancer mortality benefit (clinical-public health utility) for application in “routine-mainstream” BC probands would include the following genes: *BRCA1, BRCA2, PALB2, TP53* (under age 40), *RAD51C, RAD51D* and *BRIP1*.

Evaluated in regard of clinical-public health utility (evidenced impact on cancer mortality), the expert group recommended against inclusion on the BC-MGPT of *ATM, CHEK2, BARD1, PTEN, NF1, CDH1* and *STK11* (see Appendix Table 3 for additional WG discussion).

The only additional germline genetic testing recommended by the expert group in the diagnostic BC pathway would be for the very small proportion of BC cases actively exhibiting features strongly suggestive of a one of the rare syndromic genes, and only after (non-urgent) referral and detailed review within clinical genetics. Namely testing of *PTEN, NF1* or *STK11* where relevant syndromic manifestations of disease were present in the proband and of *CDH1* and *TP53* (in women over 40) where a strong history of relevant canonical cancers is present in the family or patient.

## Discussion

The expert group members were agreed that genetic testing in patients with BC (i) was likely to expand in volume (ii) will increasingly be performed in the mainstream oncology setting and (iii) requires delivery in time-sensitive fashion to inform immediate surgical and systemic oncological management. The expert group agreed that within mainstream oncology it is already becoming infeasible to implement at scale complex protocols involving (i) selection of genes for testing based on detailed family history scoring rules (ii) algorithmic sequential gene testing (iii) time-sensitive assessment within clinical genetics ahead of testing. Such time-consuming complexities have incentivised clinicians to default to larger pan-gene panels. This strategy obviates clinician concerns around “missing something” present within the family history (or that might subsequently emerge) and is a particularly attractive (i) in settings of ‘single-shot reimbursement’ for genetic testing, (ii) regarding potential litigiousness for failed adherence to society recommendations (iii) to seemingly offer a better “value-for-money” service.

The expert group recognised however that many mainstream oncologists (physicians/surgeons) managing patients with BC and ordering these upfront panel tests would typically have minimal experience in the downstream management of the unaffected familial GPV carriers. These oncologists therefore might not appreciate the management complexities regarding uncertainty of cancer risks, consequent management quandaries, long-term (lifelong) surveillance, reproductive decisions and concomitant potential psychological issues for GPV carriers in particular for syndromic genes.

Using panels of increasingly large numbers of genes, alongside testing of these large panels in an increasing proportion of BC probands, and then cascading out to their family members, will have exponential implications for the proportion of the population assigned as being a GPV carrier. Given the intensive protocols of decades-long, multi-organ-surveillance typically triggered by assignment of individuals as GPV-carriers, there are also substantial considerations regarding healthcare costs and resource allocation.

These issues would largely be addressed by our recommendations for inclusion on the upfront BC-MGPT of only those genes offering evidenced clinical public-health utility (cancer mortality benefit). The expert group has delineated a seven-gene BC-MGPT based on clinical public-health utility, which we propose as appropriate and sufficient for >99% of “routine-mainstream” BC cases (noting simple binary inclusion/exclusion of *TP53* based on age at BC diagnosis).

This approach reflects full consensus of the expert group against testing of rare ‘syndromic’ BCSGs in “routine-mainstream” BC cases, on the basis that identifying GPVs in this context offers minimal clinical-public health utility and likely is of net harm. The expert group agreed rare ‘syndromic’ BCSGs should only be tested where strong active indicators of a syndromic diagnosis existed. This small group of BC cases could be identified via simple questionnaire and referred for clinical genetics review downstream of their routine upfront BC-MGPT. Given that across BRIDGES, CARRIERS and UKB, the PV frequency for *TP53, PTEN, NF1, STK11* and *CDH1* combined is <0.2% (1 in 520), the proportion of BC cases requiring referral for assessment will likely be less than 1%.

*BRCA1* and *BRCA2* were agreed unanimously by the expert group as being of very high clinical-public health utility (driven by the OC risk). Scoring for *PALB2* was lower, but still attaining a mean rating of 3.44/5 (high), on the basis of TNBC association, moderate OC risk and possible therapeutic potential. There was greater debate within the expert group regarding inclusion of *TP53*, even given the pre-stated caveat of restricting testing to women age <40 years. Concerns related largely to uncertainty for the penetrance for rare cancers (especially paediatric) even in this context of ascertainment. Several expert group members advocated for recommending clinical genetics review ahead of any testing of *TP53;* counter-weighing were concerns that consequent pathway complexity would impede expedient mainstream testing at time of BC diagnosis.

*RAD51C* and *RAD51D* each attained mean scores for clinical-public health utility within the moderate range (2.17). The expert group equivocated over their inclusion on the BC-MGPT, recognising the weak overall BC association (OR<2) and very low detection rate of GPVs in unselected BC. However, with focus on clinical-public health utility, the expert group was motivated by the opportunity for OC prevention; albeit that evidence of mortality benefit was extrapolated from studies in *BRCA1/BRCA2* GPV-carriers rather than directly observed. Also factored into this assessment was the low morbidity of RRBSO in post-menopausal women, modest numbers of GPV-carriers and lack of equivocal additional cancer associations for these genes. As a logical sequel to inclusion of *RAD51C* and *RAD51D*, the expert group noted *BRIP1* as having a very similar risk profile for high-grade serous ovarian cancer to *RAD51C*/*RAD51D*. Recognising its absence of significant association with BC, the expert group saw net merit in inclusion of *BRIP1* on the BC-MGPT (thus reflecting how breast and ovarian cancers are tightly interwoven in regard of genetic risk assessment). Furthermore, the expert group members noted from recent data (and their own experience), a negligible frequency of MMR gene GPVs in unselected ovarian cancers (i.e., unselected for personal/family features of Lynch Syndrome). Accordingly, the expert group argued that the proposed BC-MGPT of *BRCA1, BRCA2, PALB2, RAD51C, RAD51D* and *BRIP1* (with *TP53* duly excluded) could equivalently serve as an ovarian cancer susceptibility panel.

Notably excluded from our BC-MGPT (constituted for clinical public health utility with a focus on impact on cancer mortality), are *CHEK2* and *ATM. CHEK2/ATM* results have in recent years been incorporated within the surgical consultation for “explaining” (in part) why the BC arose and informing the individualised contralateral risk estimation. However, it is well established that risk-reducing mastectomy based on these *CHEK2/ATM* results will not influence BC-specific mortality in the proband (the highly varied estimates for contralateral risk not withstanding)^47–49^. Whilst direct evidence from longitudinal surveillance studies in *CHEK2/ATM* PV-carriers is lacking, it is unlikely that RRBM or regimens of additional surveillance (sometimes including MRI) for unaffected *CHEK2*/*ATM* GPV carriers add significant absolute breast cancer mortality benefit over and above population-level breast screening (based on (i) limited evidence of breast cancer mortality benefit from surveillance and RRM studies in *BRCA2* PV carriers and (ii) the lower and predominantly ER-positive BC risks for *CHEK2/ATM)*^*46*^.

The expert group noted furthermore that whilst multiple associations with other cancers have been reported for *CHEK2/ATM*, there was not sizeable, reproduceable association with OC. The risk estimates have varied widely over time for other cancers and the concomitant surveillance regimens for these cancers lack evidence for net benefit improving mortality (see Appendix Tables 1,4). The expert group noted the population frequency of *CHEK2* GPVs is relatively high (even just considering the truncating variants for which breast cancer RR>2). Accordingly, it is particularly important to have demonstrated net clinical public health benefit from identification and SPED management of these individuals. In addition, there are a number of common *CHEK2* missense vars for which BC RR <1.5 (for example I157T and S428F), the widespread clinical reporting of which creates additional clinical management quandaries. Thus, the expert group judged testing these genes to offer low clinical-public health benefit overall in regard of impact on cancer mortality albeit that the results could be usefully informative for BC risk estimation (in particular as part of multifactorial risk analysis).

*BARD1* was likewise deemed of low clinical-public health benefit for inclusion on the BC-MGPT, based on its very low PV-frequency, modest BC risk and lack of association with OC.

Considerations of resource allocation are important across healthcare systems, in both high-and low-middle-income economies. Regardless of setting, the greatest system-level benefit is likely to be gained from maximising identification of GPV-carriers for the genes for which SPED interventions offer the highest mortality benefit. If, for a given gene, there is no evidence that the associated cancer risks and concomitant SPED interventions number result in improvement to cancer mortality for the GPV carriers, testing of that gene may just be causing clinical harms and diversion of healthcare resource. The economies of NGS testing has rendered the analyses of increasingly large panels of genes inevitably attractive, especially within a competitive diagnostic and clinical marketplace. However, the economic (and other) costs of downstream management of genetic findings will typically dwarf by scales of magnitude the cost of generating the, especially where decades of surveillance are initiated for multiple organ systems for multiple family members. Recognising the inevitable cautions about “putting genies back in bottles”, this incipient expansion in BC germline testing is a critical inflection point at which to reflect. Circumspection regarding clinical-public health utility, evidenced impact on mortality, avoidance of harms and health-economic benefit are of course applicable beyond patients with BC as we rapidly expand the reach of germline genetic testing.

## Data Availability

All data produced in the present work are contained in the manuscript.

## Acknowledgment

This is a project initiated by the ESMO Precision Oncology Working Group. We would like to thank ESMO leadership for their support in this manuscript. We would like to thank Sophie Allen, Institute of Cancer Research London for her assistance in preparing Table 1 and Charlie Rowlands, Institute of Cancer Research London for his assistance on collating metrics for Table 2 and Supplementary Table 3. We should like to thank Bianca Carzis for her assistance to Professor Amanda Krause in data assembly.

## Funding

This project was funded by the European Society for Medical Oncology (no grant number applies).

## Disclosures

**MIA** reports receipt of a fee for participation in Advisory Board from Pfizer; receipt of a fee as an invited speaker from AstraZeneca, Daiichi Sankio, MSD, Roche. **JB** reports receipt of a fee as an invited speaker from Astra Zeneca; educational programmes to institution from Astra Zeneca, MSD; financial interest to institution as a Steering Committee Member from Astra Zeneca; financial interest to institution as a local principal investigator from MedSir. **EC** reports receipt of a fee for participation in Advisory Board from AstraZeneca, Bayer, Daiichi-Sankyo, Janssen, Eli Lilly, Medscape, MSD, Novartis, Pfizer; receipt of a fee as an invited speaker from Astellas, AstraZeneca, Janssen, Medscape, PeerVoice, Pfizer; receipt of a fee for writing engagement from Pfizer; financial interest from receipt of research grants to institution from Bayer, Janssen, Pfizer; receipt of a fee as a Steering Committee member from Janssen, Pfizer, Telix; financial interest to institution as a local principal investigator from AstraZeneca, Janssen, Macrogenics, MSD, Pfizer. **GC** reports receipt of a fee for participation in Advisory Board from AstraZeneca, BMS, Celcuity, Daiichi Sankyo, Exact Sciences, Gilead, Eli Lilly, Menarini, Merck, Pfizer, Roche, Veracyte, Ellipsis; receipt of a fee as an invited speaker from AstraZeneca, Daiichi Sankyo, Novartis, Pfizer, Roche; receipt of a fee for writing engagement from Pfizer; receipt of funding to institution for running phase I studies from Astellas, AstraZeneca, Blueprint Medicine, BMS, Daiichi Sankyo, Kymab, Novartis, Phylogen, Roche, Sanofi; receipt of a research grant to institution for running an investigator initiated trial from Merck; receipt of a fee to institution as a coordinating principal investigator from Relay Therapeutics; non-financial interest for advisory role as an officer in Consiglio Superiore di Sanità, Italian National Health Council as an Advisor for Ministry of Health, as an Editor in Cihef of the ESMO Open, in EuropaDonna as a Member of the Scientific Council, in EUSOMA as a Member of the Advisory Council, and in Fondazione Beretta. **SD** reports receipt of a fee for providing consultancy from GSK, Intellia; no financial interest to institution as a local principal investigator from AstraZeneca. **GE** reports receipt of a fee for participation in Advisory Board from EverythingGenetic Ltd, Recursion, Springworks, Syanatra; receipt of a fee for writing engagement from AstraZeneca. **WF** reports no financial interest for a leadership role in organizing a biennial BRCA symposium which is supported by Astra Zeneca. He reports holding a research grant from Astra Zeneca. **HH** reports receipt of a fee for participation in Advisory Board from AstraZeneca; no financial interest for leadership role as a Chair of the UK Cancer Genetics Group. **NH** reports non-financial interest for leadership role in the European Commission as a Chair of the European Reference Network Genetic Tumour Risk Syndromes (ERN GENTURIS). **JN** reports non-financial interest from receipt of research grant to institution from AstraZeneca. **MR** reports receipt of a fee for participation in Advisory Board from MyMedEd; receipt of a fee as an invited speaker from Clinical Care Options, MJH Holdings, Physician’s Education Resource; receipt of a fee for review of guideline pathways from Change Healthcare; financial interest to institution as a co-principal investigator from Merck; financial interest to institution as a local principal investigator from Artios Pharma, AstraZeneca, Merck; financial interest to institution as a local co-principal investigator from Merck, Pfizer; no financial interest as a Steering Committee Member from AstraZeneca, Merck; no financial interest for advisory role from Foundation Medicine, Tempus Labs, Zenith Pharmaceuticals; no financial interest for editorial services from AstraZeneca, Merck. **CT** reports receipt of fees to charities for participation in Advisory Board from Roche and as an invited speaker from AstraZeneca. **CBW** reports receipt of a fee for participation in Advisory Board from BMS, Celgene, lncyte, Rafael, RedHill, Roche, Shire/Baxalta; receipt of a fee as an invited speaker from Amgen, AstraZeneca, Bayer, BMS, Celgene, Chugai, Falk, GSK, Janssen, Lilly, Merck, MSD, QuIP GmbH, Roche, Servier, Sirtex, Taiho; receipt of a fee for an expert testimony from Janssen; receipt of travel support from Bayer, Celgene, Janssen, RedHill, Roche, Servier, Taiho; non-financial interest from receipt of research grant both personal and to institution from Roche; non-financial interest for serving as an officer in AIO - Arbeitsgemeinschaft Internistische Onkologie (Germany); non-financial interest for advisory role in EU Commission – DG RTD as a member of the EU Commission Mission Board for Cancer, non-financial interest for advisory role in German Federal Ministry of Education and Research, Member Forum Zukunftsstrategie.

All other have declared no conflict of interest. CC, PJ, AK, KN, MT,

## References

1. Miki Y, Swensen J, Shattuck-Eidens D, et al. A strong candidate for the breast and ovarian cancer susceptibility gene BRCA1. Science (New York, NY) 1994; 266(5182): 66–71.

2. Wooster R, Bignell G, Lancaster J, et al. Identification of the breast cancer susceptibility gene BRCA2. Nature 1995; 378(6559): 789–92.

3. Smith P, McGuffog L, Easton DF, et al. A genome wide linkage search for breast cancer susceptibility genes. Genes, chromosomes & cancer 2006; 45(7): 646–55.

4. Rahman N, Seal S, Thompson D, et al. PALB2, which encodes a BRCA2-interacting protein, is a breast cancer susceptibility gene. Nature genetics 2007; 39(2): 165–7.

5. Renwick A, Thompson D, Seal S, et al. ATM mutations that cause ataxia-telangiectasia are breast cancer susceptibility alleles. Nature genetics 2006; 38(8): 873–5.

6. Meijers-Heijboer H, van den Ouweland A, Klijn J, et al. Low-penetrance susceptibility to breast cancer due to CHEK2(*)1100delC in noncarriers of BRCA1 or BRCA2 mutations. Nature genetics 2002; 31(1): 55–9.

7. Erkko H, Xia B, Nikkila J, et al. A recurrent mutation in PALB2 in Finnish cancer families. Nature 2007; 446(7133): 316–9.

8. Antoniou AC, Foulkes WD, Tischkowitz M. Breast-cancer risk in families with mutations in PALB2. The New England journal of medicine 2014; 371(17): 1651–2.

9. Dorling L, Carvalho S, Allen J, et al. Breast Cancer Risk Genes - Association Analysis in More than 113,000 Women. The New England journal of medicine 2021; 384(5): 428–39.

10. Hu C, Hart SN, Gnanaolivu R, et al. A Population-Based Study of Genes Previously Implicated in Breast Cancer. The New England journal of medicine 2021; 384(5): 440–51.

11. Rowlands CF, Allen S, Balmaña J, et al. Population-based germline breast cancer gene association studies and meta-analysis to inform wider mainstream testing. Ann Oncol 2024.

12. Heikkinen K, Rapakko K, Karppinen SM, et al. RAD50 and NBS1 are breast cancer susceptibility genes associated with genomic instability. Carcinogenesis 2006; 27(8): 1593–9.

13. Cybulski C, Carrot-Zhang J, Kluźniak W, et al. Germline RECQL mutations are associated with breast cancer susceptibility. Nature genetics 2015; 47(6): 643–6.

14. Tommiska J, Seal S, Renwick A, et al. Evaluation of RAD50 in familial breast cancer predisposition. Int J Cancer 2006; 118(11): 2911–6.

15. Landwehr R, Bogdanova NV, Antonenkova N, et al. Mutation analysis of the SLX4/FANCP gene in hereditary breast cancer. Breast cancer research and treatment 2011; 130(3): 1021–8.

16. Seal S, Thompson D, Renwick A, et al. Truncating mutations in the Fanconi anemia J gene BRIP1 are low-penetrance breast cancer susceptibility alleles. Nature genetics 2006; 38(11): 1239–41.

17. Bedrosian IA-O, Somerfield MA-O, Achatz MI, et al. Germline Testing in Patients With Breast Cancer: ASCO-Society of Surgical Oncology Guideline. (1527-7755 (Electronic)).

18. Evans DG, Howell SJ, Frayling IM, Peltonen J. Gene panel testing for breast cancer should not be used to confirm syndromic gene associations. NPJ genomic medicine 2018; 3: 32.

19. Kuchenbaecker KB, Hopper JL, Barnes DR, et al. Risks of Breast, Ovarian, and Contralateral Breast Cancer for BRCA1 and BRCA2 Mutation Carriers. Jama 2017; 317(23): 2402–16.

20. Morgan RD, Burghel GJ, Flaum N, et al. Extended panel testing in ovarian cancer reveals BRIP1 as the third most important predisposition gene. Genetics in medicine : official journal of the American College of Medical Genetics 2024; 26(10): 101230.

21. Yang X, Song H, Leslie G, et al. Ovarian and breast cancer risks associated with pathogenic variants in RAD51C and RAD51D. Journal of the National Cancer Institute 2020.

22. Norquist BM, Harrell MI, Brady MF, et al. Inherited Mutations in Women With Ovarian Carcinoma. JAMA oncology 2016; 2(4): 482–90.

23. Ramus SJ, Song H, Dicks E, et al. Germline Mutations in the BRIP1, BARD1, PALB2, and NBN Genes in Women With Ovarian Cancer. Journal of the National Cancer Institute 2015; 107(11).

24. Tischkowitz M, Balmaña J, Foulkes WD, et al. Management of individuals with germline variants in PALB2: a clinical practice resource of the American College of Medical Genetics and Genomics (ACMG). Genetics in medicine : official journal of the American College of Medical Genetics 2021; 23(8): 1416–23.

25. Mukhtar TK, Wilcox N, Dennis J, et al. Protein-truncating and rare missense variants in ATM and CHEK2 and associations with cancer in UK Biobank whole-exome sequence data. Journal of medical genetics 2024; 61(11): 1016–22.

26. Kurian AW, Ward KC, Howlader N, et al. Genetic Testing and Results in a Population-Based Cohort of Breast Cancer Patients and Ovarian Cancer Patients. Journal of clinical oncology : official journal of the American Society of Clinical Oncology 2019; 37(15): 1305–15.

27. Song H, Dicks EM, Tyrer J, et al. Population-based targeted sequencing of 54 candidate genes identifies <em>PALB2</em> as a susceptibility gene for high-grade serous ovarian cancer. Journal of medical genetics 2021; 58(5): 305–13.

28. Li S, Silvestri V, Leslie G, et al. Cancer Risks Associated With BRCA1 and BRCA2 Pathogenic Variants. Journal of clinical oncology : official journal of the American Society of Clinical Oncology 2022; 40(14): 1529–41.

29. Hu C, LaDuca H, Shimelis H, et al. Multigene Hereditary Cancer Panels Reveal High-Risk Pancreatic Cancer Susceptibility Genes. JCO precision oncology 2018; 2.

30. Hu C, Hart SN, Polley EC, et al. Association Between Inherited Germline Mutations in Cancer Predisposition Genes and Risk of Pancreatic Cancer. Jama 2018; 319(23): 2401–9.

31. Hall MJ, Bernhisel R, Hughes E, et al. Germline Pathogenic Variants in the Ataxia Telangiectasia Mutated (ATM) Gene are Associated with High and Moderate Risks for Multiple Cancers. Cancer prevention research (Philadelphia, Pa) 2021; 14(4): 433–40.

32. Yang X, Leslie G, Doroszuk A, et al. Cancer Risks Associated With Germline PALB2 Pathogenic Variants: An International Study of 524 Families. Journal of clinical oncology : official journal of the American Society of Clinical Oncology 2020; 38(7): 674–85.

33. Ryan CE, Fasaye GA, Gallanis AF, et al. Germline CDH1 Variants and Lifetime Cancer Risk. Jama 2024; 332(9): 722–9.

34. Finch AP, Lubinski J, Moller P, et al. Impact of oophorectomy on cancer incidence and mortality in women with a BRCA1 or BRCA2 mutation. Journal of clinical oncology : official journal of the American Society of Clinical Oncology 2014; 32(15): 1547–53.

35. Heemskerk-Gerritsen BA, Seynaeve C, van Asperen CJ, et al. Breast cancer risk after salpingo-oophorectomy in healthy BRCA1/2 mutation carriers: revisiting the evidence for risk reduction. Journal of the National Cancer Institute 2015; 107(5).

36. Kotsopoulos J, Lubinski J, Gronwald J, et al. Bilateral Oophorectomy and the Risk of Breast Cancer in BRCA1 Mutation Carriers: A Reappraisal. Cancer epidemiology, biomarkers & prevention : a publication of the American Association for Cancer Research, cosponsored by the American Society of Preventive Oncology 2022; 31(7): 1351–8.

37. Kotsopoulos J, Gronwald J, Huzarski T, et al. Bilateral Oophorectomy and All-Cause Mortality in Women With BRCA1 and BRCA2 Sequence Variations. JAMA oncology 2024; 10(4): 484–92.

38. Domchek SM, Friebel TM, Singer CF, et al. Association of risk-reducing surgery in BRCA1 or BRCA2 mutation carriers with cancer risk and mortality. Jama 2010; 304(9): 967–75.

39. Manchanda R, Legood R, Antoniou C, Gordeev V, Menon U. Defining the risk threshold of premenopausal risk reducing salpingo-oophorectomy for ovarian cancer prevention: a cost-effectiveness analysis. Journal of medical genetics 2016: In Press.

40. Manchanda R, Legood R, Pearce L, Menon U. Defining the risk threshold for risk reducing salpingo-oophorectomy for ovarian cancer prevention in low risk postmenopausal women. Gynecologic oncology 2015; 139(3): 487–94.

41. Lubinski J, Kotsopoulos J, Moller P, et al. MRI Surveillance and Breast Cancer Mortality in Women With BRCA1 and BRCA2 Sequence Variations. JAMA oncology 2024; 10(4): 493–9.

42. Heemskerk-Gerritsen BAM, Jager A, Koppert LB, et al. Survival after bilateral risk-reducing mastectomy in healthy BRCA1 and BRCA2 mutation carriers. Breast cancer research and treatment 2019; 177(3): 723–33.

43. Carbine NE, Lostumbo L, Wallace J, Ko H. Risk-reducing mastectomy for the prevention of primary breast cancer. The Cochrane database of systematic reviews 2018; 4(4): Cd002748.

44. Metcalfe K, Gershman S, Ghadirian P, et al. Contralateral mastectomy and survival after breast cancer in carriers of BRCA1 and BRCA2 mutations: retrospective analysis. BMJ (Clinical research ed) 2014; 348: g226.

45. Metcalfe K, Huzarski T, Gronwald J, et al. Risk-reducing mastectomy and breast cancer mortality in women with a BRCA1 or BRCA2 pathogenic variant: an international analysis. British journal of cancer 2023.

46. Lowry KP, Geuzinge HA, Stout NK, et al. Breast Cancer Screening Strategies for Women With ATM, CHEK2, and PALB2 Pathogenic Variants: A Comparative Modeling Analysis. JAMA oncology 2022; 8(4): 587–96.

47. Yadav S, Boddicker NJ, Na J, et al. Contralateral Breast Cancer Risk Among Carriers of Germline Pathogenic Variants in ATM, BRCA1, BRCA2, CHEK2, and PALB2. Journal of Clinical Oncology 2023; 41(9): 1703–13.

48. Pal T, Schon KR, Astiazaran-Symonds E, et al. Management of individuals with heterozygous germline pathogenic variants in ATM: A clinical practice resource of the American College of Medical Genetics and Genomics (ACMG). Genetics in medicine : official journal of the American College of Medical Genetics 2024: 101243.

49. Hanson H, Astiazaran-Symonds E, Amendola LM, et al. Management of individuals with germline pathogenic/likely pathogenic variants in CHEK2: A clinical practice resource of the American College of Medical Genetics and Genomics (ACMG). Genetics in medicine : official journal of the American College of Medical Genetics 2023; 25(10): 100870.

50. Owens DK, Davidson KW, Krist AH, et al. Screening for Pancreatic Cancer: US Preventive Services Task Force Reaffirmation Recommendation Statement. Jama 2019; 322(5): 438–44.

51. Grossman DC, Curry SJ, Owens DK, et al. Screening for Prostate Cancer: US Preventive Services Task Force Recommendation Statement. Jama 2018; 319(18): 1901–13.

52. Sheel ARG, Harrison S, Sarantitis I, et al. Identification of Cystic Lesions by Secondary Screening of Familial Pancreatic Cancer (FPC) Kindreds Is Not Associated with the Stratified Risk of Cancer. The American journal of gastroenterology 2019; 114(1): 155–64.

53. Dbouk M, Katona BW, Brand RE, et al. The Multicenter Cancer of Pancreas Screening Study: Impact on Stage and Survival. Journal of clinical oncology : official journal of the American Society of Clinical Oncology 2022; 40(28): 3257–66.

54. Page EC, Bancroft EK, Brook MN, et al. Interim Results from the IMPACT Study: Evidence for Prostate-specific Antigen Screening in BRCA2 Mutation Carriers. Eur Urol 2019; 76(6): 831–42.

55. Knoedler S, Jiang J, Moog P, et al. Preventive Paradox? Postoperative Outcomes After Risk-Reducing Mastectomy and Direct-to-Implant Breast Reconstruction. Clin Breast Cancer 2024.

56. Bar-Mashiah A, Soper ER, Cullina S, et al. CDH1 pathogenic variants and cancer risk in an unselected patient population. Familial cancer 2022; 21(2): 235–9.

57. Stewart DR, Frone MN, Chanock SJ. Stomaching Multigene Panel Testing: What to Do About CDH1? Journal of the National Cancer Institute 2020; 112(4): 325–6.

58. Carlsson L, Thain E, Gillies B, Metcalfe K. Psychological and health behaviour outcomes following multi-gene panel testing for hereditary breast and ovarian cancer risk: a mini-review of the literature. Hereditary cancer in clinical practice 2022; 20(1): 25.

59. Hamilton JG, Robson ME. Psychosocial Effects of Multigene Panel Testing in the Context of Cancer Genomics. Hastings Center Report 2019; 49(S1): S44–S52.

60. Hall MJ, Forman AD, Pilarski R, Wiesner G, Giri VN. Gene Panel Testing for Inherited Cancer Risk. Journal of the National Comprehensive Cancer Network J Natl Compr Canc Netw 2014; 12(9): 1339–46.

61. Domchek SM, Bradbury A, Garber JE, Offit K, Robson ME. Multiplex Genetic Testing for Cancer Susceptibility: Out on the High Wire Without a Net? Journal of Clinical Oncology 2013; 31(10): 1267–70.

62. Geyer CE, Jr., Garber JE, Gelber RD, et al. Overall survival in the OlympiA phase III trial of adjuvant olaparib in patients with germline pathogenic variants in BRCA1/2 and high-risk, early breast cancer. Ann Oncol 2022; 33(12): 1250–68.

63. Tutt ANJ, Garber JE, Kaufman B, et al. Adjuvant Olaparib for Patients with BRCA1-or BRCA2-Mutated Breast Cancer. The New England journal of medicine 2021; 384(25): 2394–405.

64. Tung NM, Robson ME, Ventz S, et al. TBCRC 048: Phase II Study of Olaparib for Metastatic Breast Cancer and Mutations in Homologous Recombination-Related Genes. Journal of clinical oncology : official journal of the American Society of Clinical Oncology 2020; 38(36): 4274–82.

65. Gruber JJ, Afghahi A, Timms K, et al. A phase II study of talazoparib monotherapy in patients with wild-type BRCA1 and BRCA2 with a mutation in other homologous recombination genes. Nat Cancer 2022; 3(10): 1181–91.

