## Supplementary Material for "Breast cancer multigene germline panel testing in mainstream oncology based on clinical-public health utility (cancer mortality benefit): ESMO Precision Oncology Working Group recommendations"

The ESMO Precision Oncology Working Group (POWG) initiated this project and assembled an international panel of experts on breast cancer genetic susceptibility to convene a sub-Working Group on breast cancer multigene panel testing (BC-MGPT), comprising clinical geneticists and medical oncologists, with individual invitations based on expertise and representation across healthcare settings.

**Meetings**

The BC-MGPT expert group convened four synchronous one-hour virtual meetings between 11/07/2024 and 3/12/2024. In between these times, the expert group communicated asynchronously via email to develop the manuscript.

**Agreed objectives to structure**

Four objectives were established by the expert group that should be addressed within the Recommendations.

1) To review genes currently included on BC-MGPTs-to identify a set of genes for review
2) To define and assemble evidence relevant for per-gene evaluations
3) To undertake individual evaluations of each gene against a range of different measures of utility
4) To integrate individual-level evaluations to provide consensus recommendations regarding genes for inclusion for a BC-MGPT applicable for “routine-mainstream” BC cases in mainstream oncology, as judged by clinical-public-health-utility (evidenced impact on cancer mortality)

**Survey to collect per-gene scorings for measures of utility**

The expert group agreed five categories against which to evaluate the utility of breast cancer susceptibility genes (BCSGs) for inclusion on a breast cancer multigene panel test (BC-MGPT) to be used universally in mainstream oncology for all women locally eligible for germline testing. Each gene was judged for its utility for each of five criteria:

1) estimation of BC risk in the proband (contralateral breast management)
2) estimation of BC risk in unaffected female relatives
3) alteration of clinical management (actionability) in the proband
4) alteration of clinical management (actionability in (unaffected) relatives
5) evidenced mortality benefit in the proband and/or relatives (clinical-public health utility)

Based on review of genes currently included on commercial and national BC-MGPTs, thirteen genes were deemed relevant for evaluation (*BRCA1, BRCA2, PALB2, CHEK2, ATM, BARD1, RAD51C, RAD51D, TP53, PTEN, NF1, STK11, CDH1*). It was established by consensus of the group that *TP53* should be scored on the basis of testing being restricted to BC cased aged <40.

Each group member submitted an individual response form comprising five scores for each of the thirteen genes. For each of the 65 gene-utility measure combinations, a mean and inter-quartile range of the 18 submissions were calculated. The mean score was used to assign to a categorical ranking of: very high (4-5), high (3-3.99 ), moderate (2-2.99), low (1-1.99),very low (0-0.99). On review of the summary results and shortlisting of genes for inclusion of the genes onto the BC-MGPT on the basis of scoring for clinical-public health utility, a fourteenth gene, BRIP1, was nominated for which the scoring process was subsequently repeated.

On review of results for the fourteen genes, the group elected for inclusion on the BC-MGPT those attaining scores of 4-5 (very high), 3-3.99 (high) and 2-2.99 (moderate)

### Supplementary Tables

***Appendix Table 1:* Impact on cancer mortality for SPED interventions for individuals at population-level risk and elevated genetic risk: ESMO Precision Oncology Working Group germline expert group appraisal of current published evidence.**

**OVARIAN CANCER
a. Risk-reducing bilateral salpingo-oophrectomy (high penetrance genes, intermediate penetrance genes)**
The expert group noted multiple longitudinal analyses of RRBSO in *BRCA1* and *BRCA2* GPV-carriers reporting ovarian cancer-specific mortality benefit, with some studies also reporting benefit on breast-cancer-specific mortality and all-cause mortality ^1-3^. The expert group noted as relevant to these data the differing risks to age 80 for OC (41% for *BRCA1*, versus 17% for *BRCA2*)^4^. The expert group noted lack of longitudinal outcome data quantifying the benefits of for RRBSO in the context of GPVs in other HBOPP genes, namely *RAD51C*, *RAD51D* and *BRIP1* for which relative risk of TOC is 6-8 and cumulative risk to age 80 is in the region of 11-13% ^5-7^. The expert group noted modelling analyses in which it was estimated that the threshold of lifetime risk of OC above which BRRSO would offer mortality benefit was estimated to be 5% ^8, 9^. In recent health economic analyses, RRBSO at age 45 has been calculated to be cost effective for those with GPVs in *RAD51C*, *RAD51D* and *BRIP1*^10^.
The expert group noted that studies of surveillance for ovarian cancer, such as PLCO, UK-FOCs and UK-TOCs, have demonstrated absence of benefit for ovarian cancer-specific mortality ^11-13^. The expert group noted that dynamic individualised CA-125 surveillance using protocols such as the Risk of Ovarian Cancer Algorithm (ROCA) likewise failed to improve cancer-specific mortality (although stage-shift was reported)^14^
***For RRBSO in females at elevated genetic risk, the expert group recognised high and well-evidenced clinical-public-health utility (cancer mortality benefit) for BRCA1 and BRCA2 GPV-carriers, with utility likely extending down to genes conferring high-intermediate risks of ovarian cancer (RAD51C, RAD51D, BRIP1).***

**BREAST CANCER
b. Breast cancer mammographic screening (population-based)**

The expert group noted that (i) the large randomised trials of population mammographic screening in the 1970s-1990s demonstrated modest improvement in breast-cancer specific mortality in women over 50 (ii) data are mixed regarding breast-cancer specific mortality benefit for routine mammography women aged <50, with sizeable rates of false positives in this group, (iii) that evidence is lacking regarding all-cause mortality benefit from routine breast cancer screening ^15^ ^16^. The expert group noted that in last two-to-three decades subsequent, and subsequent to the major breast screening trials (i) outcomes from BC treatment have improved dramatically in particular for ER-positive and HER2-positive BCs (ii) in most high-income countries rapid diagnostic pathways have resulted in the majority of symptomatic cases being diagnosed at early stage ^17-19^. The expert group noted routine population-level mammography age 50-70 becoming widely available from the 1990s in most high-income countries, with the recommended age of initiation brought forward to age 40 in several countries (including most recently the USA)^20^.

***The expert group recognised a complex evidence landscape for population-level* BC *screening with recent* treatment*-related advances in BC outcomes reducing the ‘ceiling room’ available for mortality gain from BC surveillance.***

**c. Breast cancer surveillance/risk-reducing bilateral mastectomy: high penetrance BCSGs**

The expert group noted a complex literature based primarily on *BRCA1*/*BRCA2* GPV carriers regarding risk-reducing bilateral mastectomy (RRBM) and MRI surveillance. The expert group noted (i) many early imaging studies focused on sensitivity/specificity of various imaging modalities and (ii) follow-up from early studies primarily reporting on cancer incidences rather than mortality (iii) challenges in defining the non-intervention reference group as most women had had one intervention or the other, with high rates of cross-over to BRRM in the MRI studies (iv) lack of power to assess mortality due to low numbers of events^21-23^.

The expert group noted recent international analysis of observational data on MRI versus no MRI of 2488 female *BRCA1* and *BRCA2* GPV-carriers from 59 centres in 11 countries suggested MRI surveillance to be associated with an 80% reduction in mortality in *BRCA1* GPV-carriers ^22^. The expert group noted earlier smaller studies, for example from the Netherlands and Ontario reporting absence of observed mortality benefit ^23^. The expert group noted the Hereditary Breast and Ovarian Cancer Netherlands (HEBON) study in which MRI surveillance and BRRM were compared within the cohort, mortality was reduced in the *BRCA1* BRRM group compared to the MRI surveillance group ^24^. However, the expert group noted clear reduction in the incidence of BCs in large international collaborative analysis of BRRM in *BRCA1*/*BRCA2* GPV carriers, but absence of mortality benefit on pseudo-randomised comparison ^25^. The expert group recognised that evaluation for current cohorts regarding mortality benefit from MRI surveillance/BRRM are further complicated by temporal advances in surgical approaches, advances in MRI surveillance protocols and (again), the improvement in overall survival across BC sub-types related to advances in treatment. It is possible that with much longer term follow up, the reduction in BC incidence observed following BRRM will translate into stronger evidence of BC-specific mortality benefit. The expert group note that health economic analyses indicate BRRM in *BRCA1*/*BRCA2* GPV carriers to be economically favourable (consistent with the sizeable reduction in BC incidence)^10^.

***The expert group agreed that taken together, and recognising their limitations, available data indicate likely benefit in breast-cancer specific mortality for BRCA1 GPV carriers from each of BBRM and MRI surveillance, in fitting with disease being of younger onset, higher overall risk and more typically TNBC. The expert group note that there is less evidence of mortality benefit for either intervention for BRCA2 GPV-carriers.***

**d. Breast cancer surveillance/risk-reducing bilateral mastectomy: intermediate penetrance BCSGs (IP-BCSGs)**
The expert group noted surveillance regimens for carriers of GPVs in IP-BCSGs such as *CHEK2* and *ATM* to vary widely (predicated in large part on what level of breast-screening is offered at population-level). For example, in the UK for IP-BCSG GPV-carriers, annual mammograms are offered for the decade of 40-49 (whereas those at population-level risk are offered 3-yearly mammography from age 50). In the USA, where the routine population-level USPSTF recommendation is for 2-yearly mammography from age 40, the NCCN has recommended consideration of annual MRI (with and without contrast) starting at age 30–35 years ^26^.
The expert group noted absence of patient data demonstrating mortality benefit for breast surveillance regimens for women at elevated risk and/or IP-BCSG GPV-carriers. The expert group noted the modelling study from CISNET in which a very modest absolute survival benefit for breast surveillance in GPV-carriers for IP-BCSGs was predicted ^27^.

Implementation of multifactorial risk analysis would allow identification of a small subset of CHEK2/ATM PV-carriers for whom their PRS and/or mammographic density would place them at high risk (>50% lifetime) of ER-positive BC; whilst unlikely to significant impact mortality, early detection or preventative surgery may offer mitigation of cancer-related morbidity.
***Given that (i) the data indicate equivocal survival benefit from RRM or MRI surveillance in BRCA2 GPV-carriers in whom BC risks are higher, disease is of younger onset and there is enrichment for TNBC (ii) IP-BCSGs ATM and CHEK2 are associated with more modest BC risks and near-exclusive association with ER-positive BC (iii) equivocal data regarding benefit of breast screening below 50 (iv) population-level breast screening offerings, taken together the expert group considered that the mortality benefit was likely to be negligible for enhanced surveillance or BRRM for IP-BCSGs GPV-carriers***.

**PROSTATE CANCER
e. Prostate cancer surveillance (population-based, groups at elevated genetic risk)**
The expert group noted long-standing debate regarding prostate cancer screening in regard of (i) high prevalence of indolent prostate cancer with consequent implications for overdiagnosis and overtreatment (ii) poor sensitivity-for-specificity of PSA as a screening test. The expert group noted that national bodies currently recommend against PSA prostate cancer screening (eg (i) USPSTF assignation as class C/D^28^ and (ii)UK National Screening Committee (UK-NSC) conclusion that “Screening for prostate cancer is currently not recommended in the UK”). The expert group noted that that both USPTF and UK-NSC have a caveat position of permitting testing on an individual basis following physician discussion. Initial data suggest that sensitivity for specificity might be improved with incorporation of MRI (possibly involving combination with PSA); various studies have been initiated but long-term follow-up of randomised data will be required to assess whether these protocols afford improved differentiation of lethal from indolent disease (and thus mitigating rates of overdiagnosis and overtreatment)^29-33^.

The expert group noted that association with prostate cancer has been most consistently reported for *BRCA2* with recent large prospective analyses across cancer types from CIMBA estimating association at RR = 2.22 (95% CI: 1.63 - 3.03) with no significant evidence of higher risk to younger men ^34^. In earlier prospective analyses of prostate cancers arising in *BRCA2* GPVs, the cancers were reported to be of younger onset, of higher Gleason score and of higher risk of death compared to incident population-ascertained prostate cancers, although the authors note potential ascertainment biases in these data^35^.

The expert group noted varying reports of association with prostate cancer for other HBOC genes, in particular for *ATM* and *CHEK2* (see Appendix Table 5) ^36-41^. Recent, more systematic case-control analyses suggest two-to three-fold risks, similar to *BRCA2*. For example, for *ATM*, recent case-control analysis in UKBiobank by Mukhbar et al. estimated association for protein-truncating variants (PTVs) at RR=2.35 (1.78 to 3.11), which was similar to the OR of 2.58 (95% CI 1.93–3.44) from case control analysis (adjusted for family history) by Hall et al of 677,000 individuals undertaking clinical panel testing ^42^. For *CHEK2*, in the same UKBiobank analysis by Mukhbar et al. PTVs in *CHEK2* were estimated to be associated with prostate cancer with OR=1.92 (1.59 to 2.32) ^43^.

The expert group noted that studies of prostate cancer surveillance, often combining PSA and MRI, have been initiated in GPV-carriers; nevertheless, as these studies were not randomised, it will remain highly challenging to appraise the mortality impact and extent of overdiagnosis of indolent prostate cancers in these groups of GPV-carriers (see 5c).

***The expert group agreed that further data will be required to assess net clinical-public-health utility for the prostate cancer surveillance protocols currently employed in males with germline GPVs provide.***

**PANCREATIC CANCER
f. Pancreatic cancer surveillance (groups at elevated genetic risk)**
The expert group noted overall poor survival metrics for pancreatic cancer globally, for example a five-year survival of <10% and ten year survival of <5% reported by Cancer Research UK for 2008-2018^44^. The expert group noted the lifetime risk of pancreatic cancer being ~1.7%, with an age profile of ~2/3 of cases diagnosed over the age of 70 (64.5% in UK)^44^. Accordingly, the ten-year population-level risk of pancreatic cancer for individuals aged 50-70 <0.3% (~1 in 460 in UK)^44^.

The expert group noted varying reports of association with pancreatic cancer for HBOC genes, with association most consistently reported for GPVs in *BRCA2* and *ATM* (see Appendix Table 5). The 2022 prospective analysis by CIMBA reported an overall RR for *BRCA2* of 3.34 (95% CI, 2.21 to 5.06) with evidence of higher relative risks at younger age (RR=4.92 (95% CI, 2.96 to 7.80) for age <65 years and RR> 1.77 (95% CI, 0.87 to 3.58) for age≥ 65 years) ^34^. Association of pancreatic cancer with *ATM* GPVs has also been reported in multiple studies, for example OR=4.21; 95% CI 3.24–5.47 reported by Hall et al. and RR=7.35 (4.27 to 12.66) from analysis by Muktar et al of UKBiobank ^42, 43^. Association with pancreatic cancer has been reported for *BRCA1* and *PALB2*, but less consistently.

Despite several decades of surveillance studies in individuals at elevated risk, these is still however no clear evidence for pancreatic surveillance improving cancer-specific mortality. In the EUROPAC study of pancreatic surveillance for individuals at elevated risk based on genetic findings and/or family history, a low rate of detection of pancreatic adenocarcinomas was reported across the cohort, with similar rates of lesions detected across the different risk strata ^45^. There have been recent reports that disease had been detected at earlier stage amongst non-randomised participants of the CAPS5 study undergoing pancreatic surveillance when compared to the stage distribution of disease within population registry data. Longer survival from time of detection has also been reported for those with screen-detected lesions compared to age-and sex-matched individuals within cancer registries ^46, 47^. However, it is challenging to draw conclusions from these data as to whether the surveillance confers impact on mortality: the lack of randomisation precludes robust assessment regarding the natural history of early-stage lesions, the extent of overdiagnosis and the impact of leadtime.

Based on current evidence, major national public health bodies currently recommend against screening for pancreatic cancer. For example, the USPSTF has awarded Grade D status regarding screening for pancreatic cancer in asymptomatic adults; whilst the UK-NSC also recommend against screening, stating “there is no reliable and accurate test”^48^.

***The WG agreed that further data are required to assess whether pancreatic cancer surveillance offers net clinical-public-health utility (cancer mortality benefit) for individuals with GPVs in HBOC genes (eg BRCA2, ATM).***

**RARE CANCER SUSCEPTIBILITY SYNDROMES**

**g. Multi-organ surveillance regimens for individuals with susceptibility to multiple (rare) cancers**
Numerous local, society, national and international surveillance protocols have been developed for genetic syndromes involving pleiomorphic susceptibility to multiple (often rare) cancers. These protocols typically comprise regular imaging, often using multiple modalities (MRI, CT, ultrasound) to monitor relevant organ systems for emergence of for example renal cancer, thyroid cancer, or neurological cancers (for example for *PTEN* or *NF1 GPV*-carriers); these may be combined with biochemical or cytological analyses ^49, 50^. Whole body MRI increasingly widely used, for example as part of Li Fraumeni Syndrome surveillance protocols typically in combination with separate breast MRI, brain MRI, abdominal ultrasound and/or endoscopy^51, 52^. Multi-cancer early detection (MCED) tests of blood (and urine) are being deployed within and outside of research settings, albeit that sensitivity for early-stage disease is often poor. There is particular interest in application of multi-cancer tests to those with pleiomorphic cancer susceptibility ^53^.

However, there is generally a lack of high-quality data demonstrating whether these surveillance protocols offer benefit in cancer-related mortality, or whether the earlier detection in those having surveillance is just a function of leadtime. It is of course inherently challenging to initiate randomised trials for individuals with high-risk cancer susceptibility syndromes on account of small numbers and reluctance to assign individuals to a non-intervention arm. Appraisal of the efficacy of these surveillance regimens is further confounded by uncertainty relating to overall cancer risks for GPV-carriers for these rare syndromic genes, as well as inevitable heterogeneity in risks associated with different variants and ascertainment contexts.

***The expert group noted as highly challenging to appraise whether regimens for rare cancer susceptibility syndromes provide clinical-public-health utility (cancer mortality benefit).***

**Appendix Table 2: ESMO Precision Oncology Working Group germline expert group considerations around benefit-harms trade-off from genetic testing and downstream SPED interventions**

The expert group considered the importance of recognition by both professionals and patients that any risk reducing surgery or surveillance protocol implemented in GPV-carriers constitutes a balance of physical and psychological benefits and harms. And that, in itself, receiving of a positive genetic result may also confer psychological benefits and harms.

The expert group agreed that the overall benefit-harm balance was strongly predicated on (i) robustness and stability of risk information and (ii) proven efficacy of available SPED interventions. The expert group agreed that for the rare syndromic genes, the benefit-harm balance would typically be determined by the clinical context of testing (namely whether within the patient/family, there were pre-existing related cancers and/or other clinical manifestation of the syndrome).

a) **Risk-reducing surgery can have significant short- and long-morbidity**

Surgical removal of at-risk organs remains a key intervention for many cancer susceptibility syndromes, especially where there is no effective surveillance as an alternative. However, both (i) long-term morbidities and (ii) short-term surgical and anaesthetic risks vary widely between different surgeries, for example:

- Risk reducing bilateral salpingo-oophorectomy can typically be performed laparoscopically as a day-case; significant short term morbidity is rare, especially when surgery undertaken at age>45 years and/or accompanied by time-limited hormone replacement. Judicious surgical timing is critical as premature menopause (particularly under age 45) can lead to significant impact on quality of life, and long-term health consequences regarding bone health, cardiovascular risk factor and dementia^54-57^.
- Concomitant hysterectomy adds significantly to short term morbidity from RRBSO; performance of hysterectomy in the absence of evidenced increase in endometrial risk should thus be questioned.
- Bilateral risk reducing mastectomy is a more significant operation requiring more lengthy inpatient stay and recovery which is in part dependent on choice of reconstruction; common negative sequelae include surgical complications, post mastectomy chronic pain, psychosexual impact and dissatisfaction from aesthetic outcomes, which may necessitate repeat surgeries (Knoedler, Jiang et al. 2024).
- Risk reducing	gastrectomy (most commonly undertaken	in *CDH1* heterozygotes) typically causes significant lifelong gastrointestinal and nutritional sequelae. The efficacy of gastro-oesophageal surveillance for diffuse gastric cancer is uncertain, so surgery is the only intervention proven to mitigate gastric-cancer related mortality in *CDH1* GPV-carriers. However, this high-morbidity procedure will likely cause significant net harm if undertaken without there being clearly evidenced high prior risk (Stewart, Frone et al. 2020). A recent study has suggested a lifetime risk of gastric cancer as low as 10% in CHD1 heterozygotes. If confirmed, this would have a major impact on the consideration of risks, benefits and timing of gastrectomy^58^.

b) **The harms from surveillance (screening) due to false positive results, overdiagnosis and overtreatment are widely underappreciated**

For individuals with CSG-GPVs, follow-up regimens typically comprise surveillance that is frequent (often annual), undertaken from a young age, extended often for life and with multi-modal/multi-organ approaches employed, for example whole-body MRI or overlying of multiple screening technologies (for example imaging and complex biochemistry for monitoring for pheochromocytomas). The more screening tests that are performed, the higher the likelihood of false positive results. These may necessitate follow-up investigations (perhaps longitudinally) that are more invasive and/or involve more ionising radiation.

Overdiagnosis refers to identification on screening/surveillance of asymptomatic ‘cancers’ that would otherwise have manifest or caused harm to the patient during their lifetime (because the natural history of the lesions would have been to regress, remain static or grow extremely slowly)^59^. Overdiagnosis typically leads to overtreatment, with concomitant negative side-effects such as incontinence and impotence consequent from radical prostatectomy. Overdiagnosis and overtreatment are especially problematic following surveillance of organs in which indolent lesions are common, for example the prostate, thyroid and kidney^60^.

Only with a randomised controlled trials (i) can it be demonstrated whether a surveillance/screening regimen results in genuine improvement in survival (beyond lead time) and (ii) can the impact of overdiagnosis and over-treatment be quantified^61^.

c) **The psychological burden of genetic information will vary by context**

Whilst there can be a negative psychological burden associated with genetic information, this will likely vary according to the clinical context of testing, the nature of the genetic information and the downstream healthcare interventions available.

Where there is robust evidence exist regarding (i) cancer risks being high and (ii) SPED interventions being effective, identification of a GPV in a proband and family members is widely deemed as positive, for example identification of a GPV in *BRCA1* in a woman with BC where subsequent RRBSO in the sister reveals an early in situ ovarian lesion.

The psychological burden of genetic information is likely to be more problematic on identification of GPVs for which cancer risks are low and/or uncertain (especially meaning published data may change over time) and the downstream interventions of uncertain efficacy and/or incur substantive complications/side-effects ^62-64^. Recent comparative analysis demonstrated individuals with a GPV in a moderate-penetrance gene had higher levels of genetic testing-related distress compared to those with a G GPV in a high-risk gene^65^. Variants of uncertain significance are common finding in germline genetic testing, and the prevalence of VUS is related to the number of genes which are tested. VUS can create uncertainty and anxiety in patients and providers^62, 65^.

Risk-reducing surgery in cancer-free individuals has potential to incur decisional regret, particularly where there are surgical complications, poor aesthetic outcomes and/or long-term morbidity. The potential for regret may be greater in the context of the concomitant cancer risk being modest or uncertain (meaning there had been a sizeable likelihood that the cancer may never have arisen). In addition to the issue of overdiagnosis and overtreatment, regimens of surveillance impose significant medicalisation upon well, cancer-free family-members, as well as financial costs, opportunity costs and the recurrent anxiety of awaiting surveillance results.

The psychological impact of returning genetic information will also be influenced by the familial experience of the individual. Returning of a GPV in a ‘syndromic’ BCSG (such as *CDH1* or *NF1*) to a family in which there are already multiple relevant cases of severe, young onset cancers is quite different to this result coming “out-of-the-blue” into a family in which testing was performed in an unremarkable isolated case of routine BC^66^.

d) **For testing ‘syndromic’ genes in breast cancer cases, the phenotypic context influences cancer risks and net benefit-harm**

For ‘syndromic’ BCSGs, metrics for BC risk were until recently derived from analyses within families ascertained on the basis of having multiple cases of rare/young-onset/severe disease. Recent analyses in population-ascertained BC series have revealed significantly lower risks for the ‘syndromic’ BCSGs. In part this may reflect mitigation of recruitment biases and methodological issues inherent to familial risk analyses ^67^. It may also reflect underrepresentation of extreme, young-onset, syndromic and lethal disease within population cancer cohorts.

Manifestation of extreme phenotypes and/or multiplex familial disease also likely reflect familial clustering of polygenic and/or non-genetic factors, as well as selection for specific variants of higher variant-specific risks, with concomitant elevation of risk to unaffected carriers of the familial GPV. Empirical analyses are consistent with this hypothesis, illustrating substantially higher prospective cancer risks in the context of ascertainment with a relevant strong family history than in individuals ascertained by population testing, as recently illustrated for *CDH1* ^58^.

Overall, for most syndromic BCSGs, we lack robust prospective estimates for the risk of other associated cancers in particular for GPVs ascertained away from the classical syndromic/familial presentation (for example in “routine-mainstream” cases of BC). In such contexts, not only are we uncertain as to the cancer risks appropriate to communicate to the patient, the benefit-harm balance is shifted regarding interventions such as risk-reducing surgery or surveillance, thus potentially increasing the likelihood of psychological harms.

**Appendix Table 3: ESMO Precision Oncology Working Group germline expert group considerations for testing of 14 genes against specified parameters of utility and arguments for inclusion on a BC-MGPT for clinical-public-health utility.**

The expert group assignations of utility correspond to classifications from mean scores in the WG poll [4-5 (very high), 3-3.99 (high), 2-2.99 (moderate), 1-1.99 (low), 0-0.99 (very low)]

a) ***BRCA1*, *BRCA2***

The expert group noted that for *BRCA1* and *BRCA2* the combination of (i) the magnitude of BC risk and enriched association with TNBC (in particular of *BRCA1*),(ii) the strong association with HGSOC (in particular of *BRCA1*) and (iii) the evidenced survival benefit in stratified systemic BC oncological management placed these genes in a distinct stratum compared to all other BCSGs.

BRCA1 and *BRCA2* were evaluated by the WG to be of **very high** impact across all categories of utility, including net clinical-public health utility.

b) ***PALB2***

The expert group noted the high-risk association of *PALB2* GPVs with unselected (population-type) BC of OR= 4.30 (95%CI 3.68-5.03, Table 1), with some enriched association with TNBC^68, 69^. Given the BC risk and enrichment for TNBC are broadly similar to those of BRCA2, it was deemed that the mortality impact from BRRM and MRI surveillance will be likewise similar ^70^.

The expert group also noted clinical trial data supporting clinical benefit in stratified systemic BC oncological management^70-72^. The expert group noted that association of *PALB2* GPVs with ovarian cancer of OR 2-4 has been variably reported, thus equating to a risk to age 80 of 3-6%^5^. The expert group noted reported analyses of *PALB2* GPVs in other cancers, with pancreatic and male BC most consistently associated (see Appendix Table 5b)^70, 73^.

*PALB2* was scored by the WG as being of **very** **high** impact for informing BC risk estimation and for altering clinical management (in both probands and relatives), and was evaluated to provide **high** net clinical-public health utility (evidenced cancer mortality benefit).

c) ***TP53***

The expert group noted the low frequency of *TP53* GPVs in the unselected (population-based) BC series (BRIDGES, CARRIERS, UK Biobank) (1 in 1844, see Table 1) ^69^. This observation was recognised by the expert group to be consistent with BCs in *TP53* GPV-carriers more typically occurring at very young age (<40 and even <30) and thus potentially being under-represented in population-based BC series^74^. Elevated age-specific risks for early BCs are supported by recent demonstration of a 47.7% (23.9%-64.0%) deficit of *TP53*-PVs in females compared to males in UK Biobank (a cohort that only recruited individuals aged 40-69), a figure thus arguably reflective of the penetrance for early-onset female-specific cancers in female *TP53*-GPV carriers ^75^. Thus, it was deemed by the WG that the mortality benefits for BRRM and MRI for female *BRCA1* GPV-carriers may well be extensible to *TP53*. There may in addition be mortality benefit from whole body MRI on risk of other cancers, although no randomised studies have been performed^52, 76^.

There was universal expert group agreement that testing of *TP53* should be restricted to BCs of young onset, with debate around a threshold of <40, <35 or even <30 years. Even in the context of restricting testing to young-onset BC, there remained concern within the WG regarding uncertainty for cancer risks in GPV-positive families ascertained in the absence of family history, in particular regarding childhood cancers (see Appendix Table 3d). There was related debate as to whether *TP53* testing should advised only following consultation with Clinical Genetics. Whilst this was agreed to be a preferable pathway, it was deemed infeasible in the context of dramatically-expanded mainstream testing and was agreed that net-benefit would be achieved by mainstream upfront testing in young women.

An additional and important factor in this decision was the strong corelation of Clonal Haematopoiesis of indeterminate potential (CHIP) with advancing age. *TP53* is particularly subject to being detected at low level in blood or saliva due to CHIP in older people and after chemotherapy or radiotherapy^77, 78^. There are substantial diagnostic difficulties in distinguishing CHIP from tissue mosaicism, which then necessitates obtaining multiple further clinical samples; guidelines thus largely advocate against testing *TP53* at older ages^79, 80^.

Scored specifically regarding the context of testing *TP53* in BC cases presenting at age <40, identifying a *TP53* GPV was judged by the WG to be of **very high** impact for informing BC risk estimation and for altering clinical management (in both probands and relatives), and to provide **high** net clinical-public health utility (evidenced cancer mortality benefit).

d) ***CHEK2***

The expert group noted the ~2-fold BC risk for *CHEK2* GPVs, noting GPVs to be near wholly associated with ER+ disease^69, 81, 82^.

The expert group noted that *CHEK2* status was used for contralateral BC risk estimation for the proband but that estimates for contralateral risk have varied widely^83, 84^. Nevertheless, in particular given the association is near exclusively with ER+ disease, ipsi- or contralateral breast management on the basis of *CHEK2* status is likely to be of negligible impact on cancer-specific mortality for the proband.^85^

The expert group agreed that a *CHEK2* GPV can be informative to BC risk assessment in the unaffected female relative (especially when combined with other risk factors, family history and PRS in multimodal risk assessment)^86^. However, the expert group noted that in the context of a family history of BC, presence/absence of the familial *CHEK2* GPV often did not dichotomise risk sufficient to alter management beyond that dictated on the basis of family history. The impact on management determined by the combination of *CHEK2* status and family history was noted to depend on local risk thresholds by which MRI screening was implemented^85^.

The expert group concurred that, on the basis of prospective data from *BRCA1*/*BRCA2* GPV-carriers, the impact on BC mortality in *CHEK2* GPV-carriers from enhanced surveillance protocols (MRI or mammograms) or RRM was likely to be very modest (see Appendix Table 2d). The expert group noted that multifactorial risk analysis offered opportunity for identification of a subset of *CHEK2*/ATM PV-carriers for whom their PRS and/or mammographic density would place them at very high (>50% lifetime) risk of ER-positive BC; enhanced surveillance or preventive surgery might in this group offer significant mitigation of cancer-related morbidity, even if not impacting on mortality.

The expert group also noted for *CHEK2* the multiple reported associations of modest magnitude with other cancers , albeit with inconsistency between studies and over time ^85^. For example, colorectal cancer surveillance was recommended by the NCCN on the basis of initial reported associations of *CHEK2* c.1100delC with colorectal cancer (OR=1.8 (95% CI: 1.2–2.7) and OR=1.88 (95% CI: 1.29–2.73)^87-89^; this recommendation was subsequently reversed on the basis of subsequent large retrospective studies demonstrating absence of significant association of CHEKs GPVs with colorectal cancer (see Appendix Table 5d) ^90, 91^.

The expert group also noted missense variants in *CHEK2* to be problematic; whilst for most there is insufficient data to quantify the magnitude of association with BC, for the more common missense variants (for example p.I157T, p.S428F, and p.T476M), the magnitude of association with BC lies substantially below OR=2^82, 86, 90, 91^.

The expert group noted that GPVs in *CHEK2* were comparatively frequent (1 in 73 in unselected (population-type) BC, 1 in 172 in controls, see Table 1). The expert group noted that this comparatively high frequency necessitated particularly considered scrutiny as to clinical public health utility on account of the projected detection rates in probands, with consequent volumes of cascade-identified relatives.
*CHEK2* was scored by the WG as being of **moderate/high** impact for informing BC risk estimation (in probands and relatives respectively) and **moderate** for impact regarding alteration in clinical management (in both probands and relatives). But was evaluated to provide **low** net clinical-public health utility (evidenced cancer mortality benefit).

e) ***ATM***

The expert group noted the moderate BC risk for *ATM* GPVs, again near wholly associated with ER+ disease, with broadly similar implications as for *CHEK2* in both the proband and relatives regarding both for BC risk assessment and impact upon BC mortality for the proband and female relative ^92^. The expert group noted additional contention as compared to *CHEK2* in regard of the degree to which *ATM* GPV conferred increased contralateral BC in the proband^84^.

Similar to *CHEK2*, *ATM* missense variants are problematic: as a group they are likely to have lower BC risks than PTVs and for most there is insufficient data by which to quantify the magnitude of association^43^. The exception to this is *ATM* c.7271T>G (p.Val2424Gly), for which early segregation studies and subsequent case control analyses have demonstrated higher BC risks than for PTVs ^93, 94^.

As for *CHEK2*, the modest magnitudes and wide variability over time of published risk estimates for associations of *ATM* with multiple cancers have resulted in variably extensive and frequently changing screening recommendations for *ATM* GPV-carriers. For example, the NCCN has recently expanded their recommendations for ATM PV-carriers for breast and prostate surveillance to now also include annual screening from age 50 for pancreatic cancer using contrast-enhanced magnetic resonance cholangiopancreatography (MRCP) or endoscopic ultrasound (EUS) or both^26^.

The frequency of GPVs in *ATM* GPVs was noted by the expert group as 1 in 132 in unselected (population-type) BC and 1 in 287 in controls, see Table 1), being thus lower than for *CHEK2* but still sizeable with regard to projected rates of detection in probands and family members.

*ATM* was scored by the expert group as being of **moderate** impact for informing BC risk estimation (in probands and relatives respectively) and **low**/**moderate** for impact regarding alteration in clinical management (in both probands and relatives). But was evaluated to provide **low** net clinical-public health utility (evidenced cancer mortality benefit).

f) ***BARD1***

The expert group noted the modest odds ratio (OR=2.34 (1.85-2.97) in conjunction with low frequency of GPVs in unselected BCs (1 in 672). The WG noted enrichment for association with TNBC (OR=6.26 (3.57- 10.99)), but that GPVs were still infrequent in women with TNBC (1 in 239). The WG noted absence of reproduceable associations with other cancers, notably ovarian cancer.

The expert group judged *BARD1* to be of **low/moderate** impact for informing BC risk estimation (in probands/female relatives respectively), of low impact with regarding to alteration in clinical management. *BARD1* was evaluated to provide **low** net clinical-public health utility (evidenced cancer mortality benefit).

g) ***RAD51C*, *RAD51D***

The expert group noted the low frequency of GPVs in unselected BCs (*RAD51C*: 1 in 913, *RAD51D* 1 in 1079, see Table 1) and modest-sized BC association in this context (*RAD51C*: OR= 1.53 (1.15-2.04)) and *RAD51D*: OR=1.76 (1.29-2.41), see Table 1). The expert group noted enriched association with TNBC, but that GPVs were still infrequent in TNBC (1 in 307 for *RAD51C*, 1 in 430 for *RAD51D*).

The expert group noted the well-established ~6-8 fold risk of GPVs in *RAD51C*/*RAD51D* with HGSOC, corresponding to lifetime OC risks of ~8-13% (see Appendix Table 5), for which RRBSO would be safely predicted impactful.

The expert group judged *RAD51C* and *RAD51D* to be of **low/moderate** impact for informing BC risk estimation (in probands/female relatives respectively), of **high** impact with regarding to alteration in clinical management. *RAD51C* and *RAD51D* were evaluated to provide **moderate** net clinical-public health utility (evidenced cancer mortality benefit).

h) ***CDH1***

The expert group noted the very low GPV frequency for *CDH1* in unselected BCs (1 in 2688). The expert group noted also that *CDH1* exemplified the problematic paradigm of uncertain risks and problematic benefit-risk balance for GPVs ascertained in the absence of the relevant syndromic multi-system phenotypic features. The expert group noted the recent analysis of risk of diffuse gastric cancer associated with *CDH1* GPV stratified by mechanism of ascertainment: this risk was estimated to be ~10% in the absence of family history^58, 95^.

The expert group judged *CDH1* as **moderate** impact for informing BC risk estimation, of **high** impact regarding alteration in clinical management but **low** regarding evidenced impact on cancer mortality for GPVs ascertained in the context of routine-mainstream BC.

The expert group recommended that testing for *CDH1* (i) should be restricted to the very small proportion of BC cases in which BC histology is consistent AND other characteristic syndromic features are present in the proband OR a strong family history of gastric cancer has been documented and verified (ii) required patient review by clinical genetics (iii) need not be undertaken urgently at BC diagnosis.

i) ***NF1*, *STK11***

The expert group noted the GPV frequency in unselected BCs as low for *NF1* (1 in 1,470) and very low for *STK11* (1 in 11,525). The expert group noted that for both these genes, the pathognomonic dermatological features are typically penetrant from young age. The expert group noted there to be substantial uncertainty regarding cancer risks, especially for *STK11*, and thus also uncertainty regarding the benefit-harm balance of implementation of surveillance for a GPV when ascertained in the absence of the relevant syndromic phenotypic features.

*NF1* is also subject to being detected at low level in blood in older probands due to CHIP (similar to *TP53*)^96^.

Considering *NF1* and *STK11* in the context of GPVs identified on testing in “routine-mainstream” BC, the mean of score was **low** for informing cancer risk estimation and **moderate** with regard to impact regarding alteration in clinical management. In the context of testing in “routine-mainstream” BC, *NF1* and *STK11* scored as **very low** and **low** respectively regarding evidenced impact on cancer mortality.

The expert group recommended that testing for *NF1*, *STK11* (i) should be restricted to the very small number of BC cases in which there are other characteristic syndromic features present in the proband (+/- relevant familial manifestations) (ii) required patient review by clinical genetics (iii) need not be undertaken urgently at BC diagnosis.

j) ***PTEN***

The expert group noted the very low frequency of *PTEN* GPVs in unselected (population-type) BCs (1 in 3,755). The expert group also noted, as for *STK11* and *NF1*, the uncertainty regarding risks and management when a *PTEN* GPV was ascertained in the absence of the relevant syndromic multi-system phenotypic features (ie macrocephaly, dermatological features)^97^.

*PTEN* was judged overall to be of **moderate** impact for informing cancer risk estimation, of **high** impact regarding alteration in clinical management but scored the gene as **low** regarding evidenced impact on cancer mortality when tested in the context of routine-mainstream BC.

The expert group recommended that testing for *PTEN* (i) should be restricted to the very small number of BC cases in which there are other characteristic syndromic features present in the proband (+/- relevant familial manifestations) (ii) required patient review by clinical genetics (iii) need not be undertaken urgently at BC diagnosis

k) ***BRIP1***

The expert group did not originally include *BRIP1* in its review but undertook evaluation as part of the expert group evolution.

The expert group noted that *BRIP1* as having been originally reported as a BC susceptibility gene, but with subsequent analyses refuting this association (or finding only marginal subtype specific association with triple-negative disease)^98, 99^. Notably, association for BRIP1 was non-significant for all BC in each of BRIDGES, CARRIERS and UK Biobank for all BC and in BRIDGES and CARRIERS for TNBC. On meta-analysis across the three studies, with correction for multiple testing, association for BRIP1 was non-significant for all BC (OR=1.22 (1.01-1.47), p=0.04) (and also for TNBC).

Association of *BRIP1* with ovarian cancer is however well-established and reproduceable, with a magnitude of association of OR=~9 and rate of GPVs in unselected ovarian cancer substantially greater than RAC51C and *RAD51D* combined^5^.

On additional polling of the expert group in regard to its potential inclusion on a gene panel for testing in “routine-mainstream” BC, *BRIP1* was judged overall to be of **very low/low** impact for informing cancer risk estimation, **high** with regard to impact regarding alteration in clinical management and scored **moderat**e regarding evidenced impact on cancer mortality.

***Founder variants***

The expert group noted the frequency of founder variants can vary markedly between populations and countries; there may be accordant justification for local variation in testing protocols dictated by common local founder variants.

**Appendix Table 4: Reference sources for cancer associations for breast cancer susceptibility genes**

Appendix Table 4 summarises non-systematic assembly of references sources considered by the ESMO Precision Oncology Working Group germline expert group relating to studies of the association of cancers with BCSGs .

| **Gene** | **Cancer Type** | **Relative risk** | **Absolute risk**  **(to age 80)** | **Reference; Notes** |
| --- | --- | --- | --- | --- |
| ***BRCA1*** | Ovary | SIR=49.6 [95%CI,40.0-  61.5] | 44% to age 80 | ^4^ Prospective international cohort study of 6036 *BRCA1* and 3820 *BRCA2* female carriers (CIMBA) |
|  | Male Breast | RR=4.30; 95% CI, 1.09 to 16.96), | 0.4% to age 80 | ^34^ Prospective cohort study of 3,184 *BRCA1* and 2,157 *BRCA2* families (CIMBA) |
|  | Pancreas | RR=2.36; 95% CI, 1.51 to 3.68), | 2.5% to age 80 | ^34^ Prospective cohort study of 3,184 *BRCA1* and 2,157 *BRCA2* families (CIMBA) |
|  | Stomach | RR=2.17; 95% CI, 1.25 to 3.77) | 1.6% (male)  0.7% (female) | ^34^ Prospective cohort study of 3,184 *BRCA1* and 2,157 *BRCA2* families (CIMBA) |
|  | melanoma | OR<2 | 2.5% to age 80 | ^100^ Prospective cohort study of 6,207 North American female *BRCA1* or *BRCA2* mutation carriers (c.f. population rate of melanoma of 1.5%) |
|  | pancreas | OR=2.58; 95% CI, 1.54-4.05). |  | ^101^ Comparison of 3030 North American pancreatic cancer cases sequenced with custom multiplex PCR-based panel compared to publicly available WES/WGS data for 123 136/53 105 in the individuals with exome sequence data in the public Genome Aggregation Database/Exome Aggregation Consortium database |
|  | pancreas | OR=2.95 (1.49-5.6) |  | ^102^ Comparison of 1,652 patients with pancreatic cancer sequenced using Ambry multigene panel testing compared to publicly available control data from Exome Aggregation Consortium and Genome Aggregation Database reference controls were assessed |
|  | Prostate | SIR=2.35 (95% CI 1.43–3.88) |  | ^35^ prospective UK cohort study of male *BRCA1* (n = 376) and *BRCA2* carriers (n = 447); Sixteen *BRCA1* and 26 *BRCA2* carriers were diagnosed with PCa during follow-up |
|  | prostate | OR= 3.9 (1.4–8.5)  6/692 (0.87) in cases versus 104/53,105 (0.22%) in controls |  | ^36^ 692 unselected men with metastatic prostate cancer; comparison to publicly available ExAC population data |
| ***BRCA2*** | Ovary | SIR=13.7 (95%CI, 9.1-20.7) | 17% to age 80 | ^4^ Prospective international cohort study of 6036 *BRCA1* and 3820 *BRCA2* female carriers (CIMBA) |
|  | Male Breast | RR=44.0; 95% CI, 21.3 - 90.9), | 3.8% to age 80 | ^34^ Prospective cohort study of 3,184 *BRCA1* and 2,157 *BRCA2* families (CIMBA) |
|  | Stomach | RR=3.69; (95% CI, 2.40 - 5.67), | 3.5% to age 80 | ^34^ Prospective cohort study of 3,184 *BRCA1* and 2,157 *BRCA2* families (CIMBA) |
|  | Prostate | RR=2.22; (95% CI, 1.63 - 3.03) | 27% to age 80 | ^34^ Prospective cohort study of 3,184 *BRCA1* and 2,157 *BRCA2* families (CIMBA) |
|  | Prostate | OR= 18.6 (13.2–25.3)  37/692 (5.35%) in cases versus 153/53,105 (0.29%) in controls |  | ^36^ 692 unselected men with metastatic prostate cancer; comparison to publicly available ExAC population data |
|  | Prostate | SIR=4.45 (95% CI 2.99–6.61) |  | ^35^ prospective UK cohort study of male *BRCA1* (n = 376) and *BRCA2* carriers (n = 447); Sixteen *BRCA1* and 26 *BRCA2* carriers were diagnosed with PCa during follow-up. |
|  | Melanoma | OR<2 | 2.3% to age 80 | ^100^ Prospective cohort study of 6,207 North American female *BRCA1* or *BRCA2* mutation carriers (c.f. population rate of melanoma of 1.5%) |
|  | Pancreas | RR=3.34; (95% CI, 2.21 - 5.06 | 2.5% to age 80 | ^34^ Some evidence of age-stratified effects: RR=4.92 (95% CI, 2.96 to 7.80) for age<65; RR>1.77 (95% CI, 0.87 to 3.58) for Age>65 years (P-heterogeneity=0.03). |
|  | pancreas | OR=9.07 (6.33-12.98) |  | ^102^ Patients with pancreatic cancer (N = 1,652) were identified from a 140,000-patient cohort undergoing multigene panel testing; comparison to EXAC control data |
|  | pancreas | OR=6.20 (95% CI, 4.62-8.17) |  | ^101^ Comparison of 3030 North American pancreatic cancer cases sequenced with custom multiplex PCR-based panel compared to publicly available WES/WGS data for 123 136/53 105 in the individuals with exome sequence data in the public Genome Aggregation Database/Exome Aggregation Consortium database |
| ***PALB2*** | Ovary | OR = 3.9 (95% CI 1.5-10.3) |  | ^5^ Panel testing of 940 unselected OC cases compared to 50,703 controls from the Breast Cancer Risk after Diagnostic Gene Sequencing study |
|  | Ovary | RR=2.91 (95% CI, 1.40 to 6.04), | 4.8% (95% CI: 2.4–9.7%) to age 80 | ^73^ analysis of 524 *PALB2*-positive families |
|  | Ovary | RR=3.01 (95% CI 1.59 to 5.68; | risk to age 80 to be 3.2% (95% CI: 1.8–5.7%). | ^103^ sequencing of the coding region of 54 candidate genes in 6385 invasive EOC cases and 6115 controls of broad European ancestry collected through the OCAC consortium (inc targeted sequencing, exome and array-based genotyping). Genes with an increased frequency of putative deleterious variants in cases versus controls were further examined in an independent set of 14 135 EOC cases and 28 655 controls from the Ovarian Cancer Association Consortium and the UK Biobank |
|  | Ovary | NS |  | ^104^ Next generation sequencing was used to identify germline mutations in the coding regions of four candidate susceptibility genes-BRIP1, BARD1, PALB2 and NBN-in 3236 invasive EOC case patients and 3431 control patients of European origin, and in 2000 unaffected high-risk women from a clinical screening trial of ovarian cancer (UKFOCSS) |
|  | Male Breast | RR=7.34; 95% CI, 1.28 to 42.18 | 1% (95% CI,  0.2% to 5%) | ^73^ analysis of 524 *PALB2*-positive families |
|  | Pancreatic | RR=2.37; 95% CI, 1.24 to 4.50; | 2–3% (95% CI: females: 1–4%;  males: 2–5%) | ^73^ analysis of 524 *PALB2*-positive families |
|  | pancreatic | OR=14.82 (8.12-26.22) |  | ^102^ Patients with pancreatic cancer (N = 1,652) were identified from a 140,000-patient cohort undergoing multigene panel testing; comparison to EXAC control data |
|  | pancreas | OR=2.33 (1.23-4.01) (N/S) |  | ^101^ Comparison of 3030 North American pancreatic cancer cases sequenced with custom multiplex PCR-based panel compared to publicly available WES/WGS data for 123 136/53 105 in the individuals with exome sequence data in the public Genome Aggregation Database/Exome Aggregation Consortium database |
| ***RAD51C*** | Ovarian cancer | RR=7.55, 95% CI 5.60 to 10.19; | 11% (95% CI 6% to 21%) | ^105^ analysis of 125 families with pathogenic variants in *RAD51C*, and 60 families with pathogenic variants in *RAD51D* |
|  | Ovarian cancer | OR = 8.3 (95% CI 3.1-23.1) |  | ^5^ Panel testing of 940 unselected OC cases compared to 50,703 controls from the Breast Cancer Risk after Diagnostic Gene Sequencing study |
| ***RAD51D*** | Ovarian cancer | RR=7.60 (95% CI 5.61 to 10.30) | 13% (95% CI =7% to 23%) | ^105^ analysis of 125 families with pathogenic variants in *RAD51C*, and 60 families with pathogenic variants in *RAD51D* |
|  | Ovarian cancer | OR = 6.5 (95% CI 2.1-19.7), |  | ^5^ Panel testing of 940 unselected OC cases compared to 50,703 controls from the Breast Cancer Risk after Diagnostic Gene Sequencing study |
| ***BRIP1*** | Ovarian cancer | (OR = 8.7, 95% CI 4.6-15.8) |  | ^5^ Panel testing of 940 unselected OC cases compared to 50,703 controls from the Breast Cancer Risk after Diagnostic Gene Sequencing study |
|  | Ovarian cancer | OR=11.22 for invasive EOC (95% CI = 3.22 to 34.10, P = 1 x 10(-4)) and OR=14.09 for high-grade serous disease (95% CI = 4.04 to 45.02, P = 2 x 10(-5)).  Familial studies: RR: 3.41 (95% CI = 2.12 to 5.54, P = 7x10(-7)). |  | ^104^ Next generation sequencing was used to identify germline mutations in the coding regions of four candidate susceptibility genes-BRIP1, BARD1, PALB2 and NBN-in 3236 invasive EOC case patients and 3431 control patients of European origin, and in 2000 unaffected high-risk women from a clinical screening trial of ovarian cancer (UKFOCSS) |
| ***ATM*** | Ovarian cancer | OR=3.20 (95% CI 1.66 to 6.16) |  | ^43^ case control analysis in UK Biobank (PTVs only) |
|  | pancreas | RR 7.35 (95% CI 4.27 to 12.66) |  | ^43^ case control analysis in UK Biobank (PTVs only) |
|  | prostate | RR=2.35 (95% CI 1.78-3.11) |  | ^43^ case control analysis in UK Biobank (PTVs only) |
|  | oesophagus | RR=3.90 (95% CI 1.85 -8.20) |  | ^43^ case control analysis in UK Biobank (PTVs only) |
|  | pancreas | OR=4.21; 95% CI 3.24–5.47 |  | ^42^ association analysis of 627,742 individuals undertaking cancer panel testing, adjusted for family history at ascertainment |
|  | prostate | OR=2.58 (95% CI 1.93–3.44) |  | ^42^ association analysis of 627,742 individuals undertaking cancer panel testing, adjusted for family history at ascertainment |
|  | Stomach | OR=2.97; 95% CI, 1.66-5.31) |  | ^42^ association analysis of 627,742 individuals undertaking cancer panel testing, adjusted for family history at ascertainment |
|  | ovary | OR=1.57; 95% CI, 1.35-1.83) |  | ^42^ association analysis of 627,742 individuals undertaking cancer panel testing, adjusted for family history at ascertainment |
|  | prostate | OR=4.4 (95% CI: 2.0-9.5) |  | ^38^ 13 PRACTICAL study groups comprising 5560 cases and 3353 controls of European ancestry. |
|  | prostate | 11/692 (1.59%) in cases  133/53,105(0.25%) in controls RR=6.3 (3.2–11.3) |  | ^36^ 692 unselected men with metastatic prostate cancer; comparison to publicly available ExAC population data |
|  | pancreas |  | 9.5% by age 80 years. | ^106^ 130 pancreatic cancer kindreds with pathogenic germline *ATM* variants |
|  | pancreas | OR=8.96 (6.12-12.98) |  | ^102^ Patients with pancreatic cancer (N = 1,652) were identified from a 140,000-patient cohort undergoing multigene panel testing; comparison to EXAC control data |
| ***ATM*** | pancreas | OR=5.71 (4.38-7.33) |  | ^101^ Comparison of 3030 North American pancreatic cancer cases sequenced with custom multiplex PCR-based panel compared to publicly available WES/WGS data for 123 136/53 105 in the individuals with exome sequence data in the public Genome Aggregation Database/Exome Aggregation Consortium database |
| ***CHEK2*** | prostate | RR=1.92 (1.59-2.32, p=1.18×10−11) |  | ^43^ case control analysis in UK Biobank (PTVs only) |
|  | kidney | RR=1.83 (1.06 to 3.18, p=0.032) |  | ^43^ case control analysis in UK Biobank (PTVs only) |
|  | oesophagus | RR= 2.13 (95% CI 1.10-4.10, p=0.025) |  | ^43^ case control analysis in UK Biobank (PTVs only) |
|  | ovary | RR=2.33 (1.37 to 3.97, p=1.73×10−3) |  | ^43^ case control analysis in UK Biobank (PTVs only) |
|  | prostate | *CHEK2* 1100delC mutation (OR 3.29; 95% CI 1.85-5.85; P = 0.00) |  | ^37^ Systematic review and metanalysis; 1100delC only |
|  | prostate | 28 (4.8%) germline *CHEK2* mutations (16 of which were unique) |  | ^39^ Analysis of CHEK2 in several groups of patients with prostate cancer, comprising 578 patients |
|  | prostate | 10/692 (1.87%) in cases  314/53,105 (0.61%) in controls  OR=3.1 (1.5–5.6) |  | ^36^ 692 unselected men with metastatic prostate cancer; comparison to publicly available ExAC population data |
|  | prostate |  |  | ^40^787 men with aggressive disease and 769 with nonaggressive disease. |
|  | colon | OR=0.99 (0.62-1.57) |  | ^43^ case control analysis in UK Biobank (PTVs only) |
|  | colorectal | OR=2.11 (95% CI: 1.41–3.16) |  | ^87^ Meta-analysis of six studies including 4194 CRC cases and 10,010 controls based on the search criteria were involved in this meta-analysis. A significant association of the *CHEK2* 1100delC variant with unselected CRC was found (OR=2.11, 95% CI=1.41–3.16, P=0.0003). |
|  | colorectal | OR=1.8 (95% CI: 1.2–2.7) |  | ^88^ *CHEK2* 1100delC; revised analysis of Xiang et al |
|  | colorectal | OR=1.88 (95% CI: 1.29–2.73 |  | ^89^*CHEK2* 1100delC |
|  | colorectal | OR=0.62, 95% CI 0.51-0.76; P < .001 |  | ^91^ frequency of cancers in 3783 individuals with with *CHEK2* PVs on MGPT compared to those in individuals with wildtype MGPT. No association for *CHEK2* 1100delC |
|  | colorectal | OR=1.10, 95% CI 0.91–1.33, p = 0.30600 |  | ^90^ Retrospective analysis of 705,797 patients receiving a laboratory multigene panel test. All *CHEK2* PVs |

1	Finch AP, Lubinski J, Moller P et al. Impact of oophorectomy on cancer incidence and mortality in women with a BRCA1 or BRCA2 mutation. Journal of clinical oncology : official journal of the American Society of Clinical Oncology 2014; 32 (15): 1547-1553.

2	Kotsopoulos J, Gronwald J, Huzarski T et al. Bilateral Oophorectomy and All-Cause Mortality in Women With BRCA1 and BRCA2 Sequence Variations. JAMA oncology 2024; 10 (4): 484-492.

3	Domchek SM, Friebel TM, Singer CF et al. Association of risk-reducing surgery in BRCA1 or BRCA2 mutation carriers with cancer risk and mortality. Jama 2010; 304 (9): 967-975.

4	Kuchenbaecker KB, Hopper JL, Barnes DR et al. Risks of Breast, Ovarian, and Contralateral Breast Cancer for BRCA1 and BRCA2 Mutation Carriers. Jama 2017; 317 (23): 2402-2416.

5	Morgan RD, Burghel GJ, Flaum N et al. Extended panel testing in ovarian cancer reveals BRIP1 as the third most important predisposition gene. Genetics in medicine : official journal of the American College of Medical Genetics 2024; 26 (10): 101230.

6	Ramus SJ, Song H, Dicks E et al. Germline Mutations in the BRIP1, BARD1, PALB2, and NBN Genes in Women With Ovarian Cancer. Journal of the National Cancer Institute 2015; 107 (11).

7	Yang X, Song H, Leslie G et al. Ovarian and breast cancer risks associated with pathogenic variants in RAD51C and RAD51D. Journal of the National Cancer Institute 2020.

8	Manchanda R, Legood R, Antoniou C et al. Defining the risk threshold of premenopausal risk reducing salpingo-oophorectomy for ovarian cancer prevention: a cost-effectiveness analysis. Journal of medical genetics 2016: In Press.

9	Manchanda R, Legood R, Pearce L et al. Defining the risk threshold for risk reducing salpingo-oophorectomy for ovarian cancer prevention in low risk postmenopausal women. Gynecologic oncology 2015; 139 (3): 487-494.

10	Wei X, Sun L, Slade E et al. Cost-Effectiveness of Gene-Specific Prevention Strategies for Ovarian and Breast Cancer. JAMA Netw Open 2024; 7 (2): e2355324.

11	Buys SS, Partridge E, Black A et al. Effect of screening on ovarian cancer mortality: the Prostate, Lung, Colorectal and Ovarian (PLCO) Cancer Screening Randomized Controlled Trial. Jama 2011; 305 (22): 2295-2303.

12	Jacobs IJ, Menon U, Ryan A et al. Ovarian cancer screening and mortality in the UK Collaborative Trial of Ovarian Cancer Screening (UKCTOCS): a randomised controlled trial. Lancet (London, England) 2016; 387 (10022): 945-956.

13	Menon U, Gentry-Maharaj A, Burnell M et al. Ovarian cancer population screening and mortality after long-term follow-up in the UK Collaborative Trial of Ovarian Cancer Screening (UKCTOCS): a randomised controlled trial. Lancet 2021; 397 (10290): 2182-2193.

14	Philpott S, Raikou M, Manchanda R et al. The avoiding late diagnosis of ovarian cancer (ALDO) project; a pilot national surveillance programme for women with pathogenic germline variants in BRCA1 and BRCA2. Journal of medical genetics 2023; 60 (5): 440-449.

15	Bretthauer M, Wieszczy P, Løberg M et al. Estimated Lifetime Gained With Cancer Screening Tests: A Meta-Analysis of Randomized Clinical Trials. JAMA Intern Med 2023; 183 (11): 1196-1203.

16	Jüni P, Zwahlen M. It is time to initiate another breast cancer screening trial. Annals of internal medicine 2014; 160 (12): 864-866.

17	Hensing W, Santa-Maria CA, Peterson LL et al. Landmark trials in the medical oncology management of early stage breast cancer. Semin Oncol 2020; 47 (5): 278-292.

18	Goddard KAB, Feuer EJ, Mandelblatt JS et al. Estimation of Cancer Deaths Averted From Prevention, Screening, and Treatment Efforts, 1975-2020. JAMA oncology 2024.

19	Duggan C, Trapani D, Ilbawi AM et al. National health system characteristics, breast cancer stage at diagnosis, and breast cancer mortality: a population-based analysis. The Lancet Oncology 2021; 22 (11): 1632-1642.

20	Task Force Issues Final Recommendation Statement on Screening for Breast Cancer: All women should be screened every other year starting at age 40; more research still needed in key areas. In: Taskforce UPS, ed.; 2024.

21	Sardanelli F, Magni V, Rossini G et al. The paradox of MRI for breast cancer screening: high-risk and dense breasts-available evidence and current practice. Insights Imaging 2024; 15 (1): 96.

22	Lubinski J, Kotsopoulos J, Moller P et al. MRI Surveillance and Breast Cancer Mortality in Women With BRCA1 and BRCA2 Sequence Variations. JAMA oncology 2024; 10 (4): 493-499.

23	Warner E, Zhu S, Plewes DB et al. Breast Cancer Mortality among Women with a BRCA1 or BRCA2 Mutation in a Magnetic Resonance Imaging Plus Mammography Screening Program. Cancers 2020; 12 (11).

24	Heemskerk-Gerritsen BAM, Jager A, Koppert LB et al. Survival after bilateral risk-reducing mastectomy in healthy BRCA1 and BRCA2 mutation carriers. Breast cancer research and treatment 2019; 177 (3): 723-733.

25	Metcalfe K, Huzarski T, Gronwald J et al. Risk-reducing mastectomy and breast cancer mortality in women with a BRCA1 or BRCA2 pathogenic variant: an international analysis. British journal of cancer 2023.

26	Daly MB. Genetic/Familial High-Risk Assessment: Breast, Ovarian, Pancreatic, and Prostate Version 2.2025. NCCN National Comprehensive Cancer Network; 2024.

27	Lowry KP, Geuzinge HA, Stout NK et al. Breast Cancer Screening Strategies for Women With ATM, CHEK2, and PALB2 Pathogenic Variants: A Comparative Modeling Analysis. JAMA oncology 2022; 8 (4): 587-596.

28	Fenton JJ, Weyrich MS, Durbin S et al. U.S. Preventive Services Task Force Evidence Syntheses, formerly Systematic Evidence Reviews. Prostate-Specific Antigen-Based Screening for Prostate Cancer: A Systematic Evidence Review for the U.S. Preventive Services Task Force, Rockville (MD): Agency for Healthcare Research and Quality (US) 2018.

29	Würnschimmel C, Chandrasekar T, Hahn L et al. MRI as a screening tool for prostate cancer: current evidence and future challenges. World J Urol 2023; 41 (4): 921-928.

30	Grossman DC, Curry SJ, Owens DK et al. Screening for Prostate Cancer: US Preventive Services Task Force Recommendation Statement. Jama 2018; 319 (18): 1901-1913.

31	Eldred-Evans D, Tam H, Sokhi H et al. Rethinking prostate cancer screening: could MRI be an alternative screening test? Nature Reviews Urology 2020; 17 (9): 526-539.

32	Padhani AR, Godtman RA, Schoots IG. Key learning on the promise and limitations of MRI in prostate cancer screening. European Radiology 2024: 1-7.

33	Hugosson J, Godtman RA, Wallstrom J et al. Results after four years of screening for prostate cancer with PSA and MRI. New England Journal of Medicine 2024; 391 (12): 1083-1095.

34	Li S, Silvestri V, Leslie G et al. Cancer Risks Associated With BRCA1 and BRCA2 Pathogenic Variants. Journal of clinical oncology : official journal of the American Society of Clinical Oncology 2022; 40 (14): 1529-1541.

35	Nyberg T, Frost D, Barrowdale D et al. Prostate Cancer Risks for Male BRCA1 and BRCA2 Mutation Carriers: A Prospective Cohort Study. Eur Urol 2020; 77 (1): 24-35.

36	Pritchard CC, Mateo J, Walsh MF et al. Inherited DNA-Repair Gene Mutations in Men with Metastatic Prostate Cancer. The New England journal of medicine 2016; 375 (5): 443-453.

37	Wang Y, Dai B, Ye D. CHEK2 mutation and risk of prostate cancer: a systematic review and meta-analysis. Int J Clin Exp Med 2015; 8 (9): 15708-15715.

38	Karlsson Q, Brook MN, Dadaev T et al. Rare Germline Variants in ATM Predispose to Prostate Cancer: A PRACTICAL Consortium Study. Eur Urol Oncol 2021; 4 (4): 570-579.

39	Dong X, Wang L, Taniguchi K et al. Mutations in CHEK2 associated with prostate cancer risk. American journal of human genetics 2003; 72 (2): 270-280.

40	Nguyen-Dumont T, MacInnis RJ, Steen JA et al. Rare germline genetic variants and risk of aggressive prostate cancer. Int J Cancer 2020; 147 (8): 2142-2149.

41	Leongamornlert D, Saunders E, Dadaev T et al. Frequent germline deleterious mutations in DNA repair genes in familial prostate cancer cases are associated with advanced disease. British journal of cancer 2014; 110 (6): 1663-1672.

42	Hall MJ, Bernhisel R, Hughes E et al. Germline Pathogenic Variants in the Ataxia Telangiectasia Mutated (ATM) Gene are Associated with High and Moderate Risks for Multiple Cancers. Cancer prevention research (Philadelphia, Pa) 2021; 14 (4): 433-440.

43	Mukhtar TK, Wilcox N, Dennis J et al. Protein-truncating and rare missense variants in ATM and CHEK2 and associations with cancer in UK Biobank whole-exome sequence data. Journal of medical genetics 2024; 61 (11): 1016-1022.

44	Huntley C, Torr B, Sud A et al. Utility of polygenic risk scores in UK cancer screening: a modelling analysis. The Lancet Oncology 2023; 24 (6): 658-668.

45	Sheel ARG, Harrison S, Sarantitis I et al. Identification of Cystic Lesions by Secondary Screening of Familial Pancreatic Cancer (FPC) Kindreds Is Not Associated with the Stratified Risk of Cancer. The American journal of gastroenterology 2019; 114 (1): 155-164.

46	Blackford AL, Canto MI, Dbouk M et al. Pancreatic Cancer Surveillance and Survival of High-Risk Individuals. JAMA oncology 2024; 10 (8): 1087-1096.

47	Dbouk M, Katona BW, Brand RE et al. The Multicenter Cancer of Pancreas Screening Study: Impact on Stage and Survival. Journal of clinical oncology : official journal of the American Society of Clinical Oncology 2022; 40 (28): 3257-3266.

48	Owens DK, Davidson KW, Krist AH et al. Screening for Pancreatic Cancer: US Preventive Services Task Force Reaffirmation Recommendation Statement. Jama 2019; 322 (5): 438-444.

49	Tischkowitz M, Colas C, Pouwels S et al. Cancer Surveillance Guideline for individuals with PTEN hamartoma tumour syndrome. European journal of human genetics : EJHG 2020; 28 (10): 1387-1393.

50	Carton C, Evans DG, Blanco I et al. ERN GENTURIS tumour surveillance guidelines for individuals with neurofibromatosis type 1. EClinicalMedicine 2023; 56: 101818.

51	Omran M, Blomqvist L, Brandberg Y et al. Whole-body MRI within a surveillance program for carriers with clinically actionable germline TP53 variants - the Swedish constitutional TP53 study SWEP53. Hereditary cancer in clinical practice 2020; 18: 1.

52	Ballinger ML, Best A, Mai PL et al. Baseline Surveillance in Li-Fraumeni Syndrome Using Whole-Body Magnetic Resonance Imaging: A Meta-analysis. JAMA oncology 2017; 3 (12): 1634-1639.

53	Klein EA, Richards D, Cohn A et al. Clinical validation of a targeted methylation-based multi-cancer early detection test using an independent validation set. Ann Oncol 2021; 32 (9): 1167-1177.

54	Georgakis MK, Beskou-Kontou T, Theodoridis I et al. Surgical menopause in association with cognitive function and risk of dementia: A systematic review and meta-analysis. Psychoneuroendocrinology 2019; 106: 9-19.

55	Rivera CM, Grossardt BR, Rhodes DJ et al. Increased cardiovascular mortality after early bilateral oophorectomy. Menopause 2009; 16 (1): 15-23.

56	Rocca WA, Grossardt BR, Miller VM et al. Premature menopause or early menopause and risk of ischemic stroke. Menopause 2012; 19 (3): 272-277.

57	Shuster LT, Gostout BS, Grossardt BR et al. Prophylactic oophorectomy in premenopausal women and long-term health. Menopause Int 2008; 14 (3): 111-116.

58	Ryan CE, Fasaye GA, Gallanis AF et al. Germline CDH1 Variants and Lifetime Cancer Risk. Jama 2024; 332 (9): 722-729.

59	Brodersen J, Schwartz LM, Heneghan C et al. Overdiagnosis: what it is and what it isn't. BMJ Evid Based Med 2018; 23 (1): 1-3.

60	Srivastava S, Koay EJ, Borowsky AD et al. Cancer overdiagnosis: a biological challenge and clinical dilemma. Nature reviews Cancer 2019; 19 (6): 349-358.

61	Kale MS, Korenstein D. Overdiagnosis in primary care: framing the problem and finding solutions. BMJ (Clinical research ed) 2018; 362: k2820.

62	Carlsson L, Thain E, Gillies B et al. Psychological and health behaviour outcomes following multi-gene panel testing for hereditary breast and ovarian cancer risk: a mini-review of the literature. Hereditary cancer in clinical practice 2022; 20 (1): 25.

63	Hamilton JG, Robson ME. Psychosocial Effects of Multigene Panel Testing in the Context of Cancer Genomics. Hastings Center Report 2019; 49 (S1): S44-S52.

64	Hall MJ, Forman AD, Pilarski R et al. Gene Panel Testing for Inherited Cancer Risk. Journal of the National Comprehensive Cancer Network J Natl Compr Canc Netw 2014; 12 (9): 1339-1346.

65	Carlsson L, Bedard PL, Kim RH et al. Psychological distress following multi-gene panel testing for hereditary breast and ovarian cancer risk. Journal of genetic counseling 2024.

66	Chaillet KS, Sleiman MM, Jr., Yockel MR et al. Effects of personal cancer history and genomic risk information on mothers' psychological adaptation to inherited breast/ovarian cancer syndrome. Psychol Health Med 2024: 1-12.

67	Easton DF, Pharoah PD, Antoniou AC et al. Gene-panel sequencing and the prediction of breast-cancer risk. The New England journal of medicine 2015; 372 (23): 2243-2257.

68	Woodward ER, van Veen EM, Evans DG. From BRCA1 to Polygenic Risk Scores: Mutation-Associated Risks in Breast Cancer-Related Genes. Breast Care (Basel) 2021; 16 (3): 202-213.

69	Rowlands CF, Allen S, Balmaña J et al. Population-based germline breast cancer gene association studies and meta-analysis to inform wider mainstream testing. Ann Oncol 2024.

70	Tischkowitz M, Balmaña J, Foulkes WD et al. Management of individuals with germline variants in PALB2: a clinical practice resource of the American College of Medical Genetics and Genomics (ACMG). Genetics in medicine : official journal of the American College of Medical Genetics 2021; 23 (8): 1416-1423.

71	Tung NM, Robson ME, Ventz S et al. TBCRC 048: Phase II Study of Olaparib for Metastatic Breast Cancer and Mutations in Homologous Recombination-Related Genes. Journal of clinical oncology : official journal of the American Society of Clinical Oncology 2020; 38 (36): 4274-4282.

72	Gruber JJ, Afghahi A, Timms K et al. A phase II study of talazoparib monotherapy in patients with wild-type BRCA1 and BRCA2 with a mutation in other homologous recombination genes. Nat Cancer 2022; 3 (10): 1181-1191.

73	Yang X, Leslie G, Doroszuk A et al. Cancer Risks Associated With Germline PALB2 Pathogenic Variants: An International Study of 524 Families. Journal of clinical oncology : official journal of the American Society of Clinical Oncology 2020; 38 (7): 674-685.

74	Evans DG, Howell SJ, Frayling IM et al. Gene panel testing for breast cancer should not be used to confirm syndromic gene associations. NPJ genomic medicine 2018; 3: 32.

75	Wade I, Witkowski L, Ahmed A et al. Using cancer phenotype sex-specificity to enable unbiased penetrance estimation of SMARCA4 pathogenic variants for small cell carcinoma of the ovary, hypercalcemic type (SCCOHT). Genetics in medicine : official journal of the American College of Medical Genetics 2024; 27 (1): 101287.

76	Villani A, Shore A, Wasserman JD et al. Biochemical and imaging surveillance in germline TP53 mutation carriers with Li-Fraumeni syndrome: 11 year follow-up of a prospective observational study. The Lancet Oncology 2016; 17 (9): 1295-1305.

77	Coffee B, Cox HC, Bernhisel R et al. A substantial proportion of apparently heterozygous TP53 pathogenic variants detected with a next-generation sequencing hereditary pan-cancer panel are acquired somatically. Human mutation 2020; 41 (1): 203-211.

78	Schwartz AN, Hyman SR, Stokes SM et al. Evaluation of TP53 Variants Detected on Peripheral Blood or Saliva Testing: Discerning Germline From Somatic TP53 Variants. JCO precision oncology 2021; 5: 1677-1686.

79	Frebourg T, Bajalica Lagercrantz S, Oliveira C et al. Guidelines for the Li-Fraumeni and heritable TP53-related cancer syndromes. European journal of human genetics : EJHG 2020; 28 (10): 1379-1386.

80	Evans DG, Woodward ER, Bajalica-Lagercrantz S et al. Germline TP53 Testing in Breast Cancers: Why, When and How? Cancers 2020; 12 (12).

81	Dorling L, Carvalho S, Allen J et al. Breast cancer risks associated with missense variants in breast cancer susceptibility genes. Genome medicine 2022; 14 (1): 51.

82	Hu C, Hart SN, Gnanaolivu R et al. A Population-Based Study of Genes Previously Implicated in Breast Cancer. The New England journal of medicine 2021; 384 (5): 440-451.

83	Morra A, Mavaddat N, Muranen TA et al. The impact of coding germline variants on contralateral breast cancer risk and survival. The American Journal of Human Genetics 2023; 110 (3): 475-486.

84	Yadav S, Boddicker NJ, Na J et al. Contralateral Breast Cancer Risk Among Carriers of Germline Pathogenic Variants in ATM, BRCA1, BRCA2, CHEK2, and PALB2. Journal of Clinical Oncology 2023; 41 (9): 1703-1713.

85	Hanson H, Astiazaran-Symonds E, Amendola LM et al. Management of individuals with germline pathogenic/likely pathogenic variants in CHEK2: A clinical practice resource of the American College of Medical Genetics and Genomics (ACMG). Genetics in medicine : official journal of the American College of Medical Genetics 2023; 25 (10): 100870.

86	Hanson H, Astiazaran-Symonds E, Amendola LM et al. Management of individuals with germline pathogenic/likely pathogenic variants in CHEK2: A clinical practice resource of the American College of Medical Genetics and Genomics (ACMG). Genetics in Medicine 2023; 25 (10): 100870.

87	Xiang H-p, Geng X-p, Ge W-w et al. Meta-analysis of CHEK2 1100delC variant and colorectal cancer susceptibility. European Journal of Cancer 2011; 47 (17): 2546-2551.

88	Katona BW, Yang Y-X. Colorectal cancer risk associated with the CHEK2 1100delC variant. European Journal of Cancer 2017; 83: 103-105.

89	Ma X, Zhang B, Zheng W. Genetic variants associated with colorectal cancer risk: comprehensive research synopsis, meta-analysis, and epidemiological evidence. Gut 2014; 63 (2): 326.

90	Mundt E, Mabey B, Rainville I et al. Breast and colorectal cancer risks among over 6,000 CHEK2 pathogenic variant carriers: A comparison of missense versus truncating variants. Cancer Genet 2023; 278-279: 84-90.

91	Bychkovsky BL, Agaoglu NB, Horton C et al. Differences in Cancer Phenotypes Among Frequent CHEK2 Variants and Implications for Clinical Care-Checking CHEK2. JAMA oncology 2022; 8 (11): 1598-1606.

92	Pal T, Schon KR, Astiazaran-Symonds E et al. Management of individuals with heterozygous germline pathogenic variants in ATM: A clinical practice resource of the American College of Medical Genetics and Genomics (ACMG). Genetics in medicine : official journal of the American College of Medical Genetics 2024: 101243.

93	Goldgar DE, Healey S, Dowty JG et al. Rare variants in the ATM gene and risk of breast cancer. Breast cancer research : BCR 2011; 13 (4): R73.

94	Southey MC, Goldgar DE, Winqvist R et al. PALB2, CHEK2 and ATM rare variants and cancer risk: data from COGS. Journal of medical genetics 2016; 53 (12): 800-811.

95	Stewart DR, Frone MN, Chanock SJ. Stomaching Multigene Panel Testing: What to Do About CDH1? Journal of the National Cancer Institute 2020; 112 (4): 325-326.

96	Safonov AM, Speare V, Dolinsky JS et al. Clinical and molecular characteristics of NF1 mutations identified on hereditary cancer multi-gene panels. Journal of Clinical Oncology 2018; 36 (15_suppl): e13612-e13612.

97	Hendricks LAJ, Hoogerbrugge N, Mensenkamp AR et al. Cancer risks by sex and variant type in PTEN hamartoma tumor syndrome. Journal of the National Cancer Institute 2023; 115 (1): 93-103.

98	Couch FJ, Shimelis H, Hu C et al. Associations Between Cancer Predisposition Testing Panel Genes and Breast Cancer. JAMA oncology 2017; 3 (9): 1190-1196.

99	Easton DF, Lesueur F, Decker B et al. No evidence that protein truncating variants in BRIP1 are associated with breast cancer risk: implications for gene panel testing. Journal of medical genetics 2016; 53 (5): 298-309.

100	Narod SA, Metcalfe K, Finch A et al. The risk of skin cancer in women who carry BRCA1 or BRCA2 mutations. Hereditary cancer in clinical practice 2024; 22 (1): 7.

101	Hu C, Hart SN, Polley EC et al. Association Between Inherited Germline Mutations in Cancer Predisposition Genes and Risk of Pancreatic Cancer. Jama 2018; 319 (23): 2401-2409.

102	Hu C, LaDuca H, Shimelis H et al. Multigene Hereditary Cancer Panels Reveal High-Risk Pancreatic Cancer Susceptibility Genes. JCO precision oncology 2018; 2.

103	Song H, Dicks EM, Tyrer J et al. Population-based targeted sequencing of 54 candidate genes identifies <em>PALB2</em> as a susceptibility gene for high-grade serous ovarian cancer. Journal of medical genetics 2021; 58 (5): 305-313.

104	Ramus SJ, Song H, Dicks E et al. Germline Mutations in the BRIP1, BARD1, PALB2, and NBN Genes in Women With Ovarian Cancer. JNCI: Journal of the National Cancer Institute 2015; 107 (11).

105	Yang X, Song H, Leslie G et al. Ovarian and Breast Cancer Risks Associated With Pathogenic Variants in RAD51C and RAD51D. JNCI: Journal of the National Cancer Institute 2020; 112 (12): 1242-1250.

106	Hsu F-C, Roberts NJ, Childs E et al. Risk of Pancreatic Cancer Among Individuals With Pathogenic Variants in the ATM Gene. JAMA oncology 2021; 7 (11): 1664-1668.
